# Detecting changes in population trends in infection surveillance using community SARS-CoV-2 prevalence as an exemplar

**DOI:** 10.1101/2022.09.14.22279931

**Authors:** Emma Pritchard, Karina-Doris Vihta, David W. Eyre, Susan Hopkins, Tim EA Peto, Philippa C. Matthews, Nicole Stoesser, Ruth Studley, Emma Rourke, Ian Diamond, Koen B. Pouwels, Ann Sarah Walker, the COVID-19 Infection Survey Team

**Author notes:** contribution considered equal (Koen B. Pouwels, Ann Sarah Walker).

## Abstract

**Background:** Monitoring infection trends is vital to informing public health strategy. Detecting and quantifying changes in growth rates can inform policymakers’ rationale for implementing or continuing interventions aimed at reducing impact. Substantial changes in SARS-CoV-2 prevalence with emergence of variants provides opportunity to investigate different methods to do this.

**Methods:** We included PCR results from all participants in the UK’s COVID-19 Infection Survey between 1 August 2020-30 June 2022. Change-points for growth rates were identified using iterative sequential regression (ISR) and second derivatives of generalised additive models (GAMs).

Consistency between methods and timeliness of detection were compared.

**Findings:** Of 8,799,079 visits, 147,278 (1·7%) were PCR-positive. Over the time period, change-points associated with emergence of major variants were estimated to occur a median 4 days earlier (IQR 0-8) in GAMs versus ISR, with only 2/48 change-points identified by only one method. Estimating recent change-points using successive data periods, four change-points (4/96) identified by GAMs were not found when adding later data or by ISR; 77% (74/96) of change-points identified by successive GAMs were identified by ISR. Change-points were detected 3-5 weeks after they occurred in both methods but could be detected earlier within specific subgroups.

**Interpretation:** Change-points in growth rates of SARS-CoV-2 can be detected in near real-time using ISR and second derivatives of GAMs. To increase certainty about changes in epidemic trajectories both methods could be run in parallel. Running either method in near real-time on different infection surveillance data streams could provide timely warnings of changing underlying epidemiology.

**Funding:** UK Health Security Agency, Department of Health and Social Care (UK), Welsh Government, Department of Health (on behalf of the Northern Ireland Government), Scottish Government, National Institute for Health Research.

## Introduction

Infectious disease surveillance has two broad goals; identifying outbreaks which lead to sudden changes in incidence/prevalence, and detecting the emergence of a more virulent/resistant strain. Methods for such “change-point” detection are widely established, but many look for abrupt changes consistent with the former, e.g. a change in mean levels in a time series, rather than the more gradual trend changes^1^ characteristic of the latter.

Two methods considering more gradual changes and finding change-points in trends are iterative sequential regression^2,3^ (ISR) and second derivatives of generalised additive models (GAMs).^4^ ISR provides a clear statistical assessment of when rates change and estimates constant growth rates between change-points, but considers data sequentially, fixing change-points as it iterates, thus not necessarily optimising overall model fit. Second derivatives of GAMs have been used to identify periods of changes,^5,6^ and quantify change-points.^4^ The flexibility afforded by GAMs allows estimates to closely reflect reality, but it is unclear to what extent smoothing through penalized splines potentially reduces the ability to detect change-points in near real-time. While both methods have been evaluated separately,^2,6^ to our knowledge, they have never been directly compared.

Rapid changes in SARS-CoV-2 prevalence coupled with emergence of multiple variants over two years of the COVID-19 pandemic provides an ideal opportunity to test these different methods. Further, many countries are reducing restrictions and ceasing widespread testing,^7^ so identifying changes in remaining large-scale surveillance monitoring of SARS-CoV-2 positivity in the UK^8,9^ and globally,^10–13^ via optimal methods for detecting trend changes would be useful for monitoring positivity in real-time, potentially prompting surge testing amongst other interventions.

We therefore tested these two change-point detection methods using the UK’s Office for National Statistics (ONS) COVID-19 Infection Survey, as an exemplar for surveillance more generally, e.g. emergence of ribotype-027 *Clostridium difficile* or increasing antimicrobial resistance in gram-negative bacteraemia. We assessed the consistency and timeliness of detection between ISR and second derivatives of GAMs for identifying changes in growth rates of SARS-CoV-2 positivity over time. We further assessed whether earlier detection was possible considering positivity separately by age group, or split by available proxies for viral variant.

## Methods

### Study design

The ONS COVID-19 Infection Survey is a large household survey with longitudinal follow-up (ISRCTN21086382). Private households are selected randomly from address lists and previous ONS surveys on a continuous basis to provide a representative sample across the UK. Following verbal consent, a study worker visited each household to take written informed consent for individuals aged ≥2 years (from parents/carers for those 2–15 years; those 10–15 years also provided written assent). The study received ethical approval from the South Central Berkshire B Research Ethics Committee (20/SC/0195). At the first visit, participants were asked for consent for optional follow-up visits every week for the next month, then monthly thereafter (>98·5% providing such consent). At each visit, participants provided a nose and throat self-swab and completed questionnaires (https://www.ndm.ox.ac.uk/covid-19/covid-19-infection-survey/case-record-forms).

### Study Population

Analysis included all visits with positive or negative swabs from 1^st^ August 2020-30th June 2022 (n=225,348 (2%) visits with void/missing results excluded).

### Statistical Analyses

Our outcome measure was the proportion of visits with PCR-positive SARS-CoV-2 tests. We compared two methods for detecting changes in trend over time: ISR^2^ and second derivatives of GAMs.^4^ All models were run separately for 12 geographical regions (9 English regions and 3 devolved administrations: Wales, Scotland, Northern Ireland) due to positivity trend differences and ISR estimating change-points in a single time-series. Running GAMs separately by region made small differences to predictions versus including region-time interactions, and reduced computational time (**Figure S1**).

ISR, using a negative-binomial distribution with log link allowing for overdispersion, initially fitted a log-linear trend within the first month of data to 1st September 2020. Three days’ data were sequentially added to the time-series, fixing change-points if a new trend reduced the AIC by ≥6.635 (critical value at p=0·01, to reduce the impact of false-positives). If a change-point was fixed, a new change-point was not considered in the subsequent seven days. Change-points, and dates change-points were permanently fixed into the model (“detection date”), were extracted from fitted models (**Supplementary Methods**).

GAMs, using a negative-binomial distribution with log link, included a single explanatory variable of time, measured in days since 1^st^ August 2020 and modelled using thin plate splines.^14^ The number of basis functions, *k*, determining smoothness, was selected from 25, 50, 75, 100 as the lowest value with predicted positivity within ±0·25% (absolute scale) compared with *k*=100, optimising computational time, without large increases in the effective degrees of freedom^15^ (**Figure S2**). Splines were penalised based on the third derivative as the second derivative was the measure of interest.

Derivatives were estimated for smooth functions using posterior simulation with a Metropolis-Hastings sampler (as implemented in the gam.mh function from the *mgcv* R package)^16,17^ as standard Gaussian approximation will be poor in periods of low positivity (details in **Supplementary Methods**). Code was adapted from the *derivatives* function in the R *gratia* package, which currently can only obtain derivatives on the linear predictor scale.^18^ Change-points were defined at the first day zero was no longer within the 95% credible interval of the second derivative, corresponding to a 97.5% probability of a change, from 1^st^ September 2020 onwards. Positivity trends over the full time-series were compared between ISR and GAMs. Change-points were classified as found by both methods if within ±7 days. Change-points corresponding to the emergence of Alpha, Delta, BA.1, and BA.2 variants were compared between methods.

As ISR fixes change-points once found and adds data progressively, it does not need to be run on segments of data sequentially to assess real-time detection. When applied in real-time, one could run ISR from the latest detected change-point onwards to decrease fitting time, albeit change-points may differ slightly versus models incorporating the full time-series as previous data can impact AIC. To assess consistency in near real-time detection between the methods, GAMs were run sequentially adopting a sliding-window approach. Sliding-window length was determined by running GAMs on shorter time periods (16, 24, and 32-weeks) and assessing whether similar change-points were found in the final 8-weeks as most recent changes are of most interest in near real-time. Starting from 1st October 2020 (including data from 1st August 2020), seven-day increments of data were added until the sliding-window length was reached, from which seven-days of data were removed from the start of the time-series each time seven-days were added on. We selected *k* as before for sliding-window length, scaling *k* down proportionally for the shorter time-series. We checked whether all change-points identified in the last 8-weeks of each model were detected within ±7 days in five subsequent models and/or by ISR. Due to long runtimes (~12-36 hours per region including derivatives estimation), we compared GAM detection dates for the largest (London) and smallest (Northern Ireland) regions. A “detection date” for change-points identified in the GAM including data from the full time-series was defined as the last date included in the earliest successive GAM which also confirmed the change-point within ±7 days.

Using the second derivative of GAMs risks potentially missing change-points if positivity decreases and increases at the same rate over a short period of time. While the second derivative will be significantly different from zero, a new change-point will not be found when positivity changes direction as the second derivative may not cross zero. We summarised the number and position of additional change-points added if placed where, over a period of the second derivative being significantly different from zero, the first derivative changed from significantly positive to negative, or visa-versa.

### Sensitivity Analysis

To assess whether earlier detection of change-points was possible by focusing on high-risk population subgroups, change-points estimated from separate ISR and GAMs in those aged 2 years (y)-school year (sy) 11 (~ aged 16y), 12sy–49y, and 50y+, were compared with combined all-age estimates. We also considered separate analysis by PCR gene positivity as a proxy for SARS-CoV-2 variant; Delta and BA.2 being spike (S) gene target positive (SGTP), whereas Alpha and BA.1 had S-gene target failure (SGTF). Models were run separately with SGTP and SGTF positivity as outcomes, with all other positives (including those positive on only the N gene or ORF1ab) in the negative comparator group, comparing change-points to the “all positives” model.

All analysis was conducted in R version 4.0.2. Key analysis code is available at https://github.com/EmmaPritchard.

### Role of the funding source

The funder had no role in study design, data collection, data analysis, data interpretation, or writing of the report. All authors had access to all data reported in the study and accept responsibility for the decision to submit for publication.

## Results

From 1^st^ August 2020-30th June 2022, 8,799,079 visits from 533,157 participants in 266,400 households returned 147,278 (1·7%) SARS-CoV-2 positive swabs (visit characteristics in **Table S1**). From August-November 2020 (pre-Alpha), positivity rose to ~1%, before increasing to ~2% in January 2021 (Alpha; **Figure 1A; Figure S3**). Positivity decreased until June 2021 before increasing to ~1-2% in July-December 2021 (Delta). Positivity rose sharply to ~6% from December 2021 (BA.1), decreasing to ~3·5% by February 2022, before increasing to ~7·5% by mid-March (BA.2). Rises in BA.4/BA.5 began June 2022. During the pre-Alpha period, 10% of strong positives (Ct<30) had SGTF, versus 79%, 1%, 84%, and 9% in Alpha, Delta, BA.1, and BA.2-dominant periods (**Table S2**). Positivity varied by region, particularly between Northern/Southern English regions e.g. higher positivity pre-Alpha in Yorkshire, versus London (**Figure S3B**).

**Figure 1:**
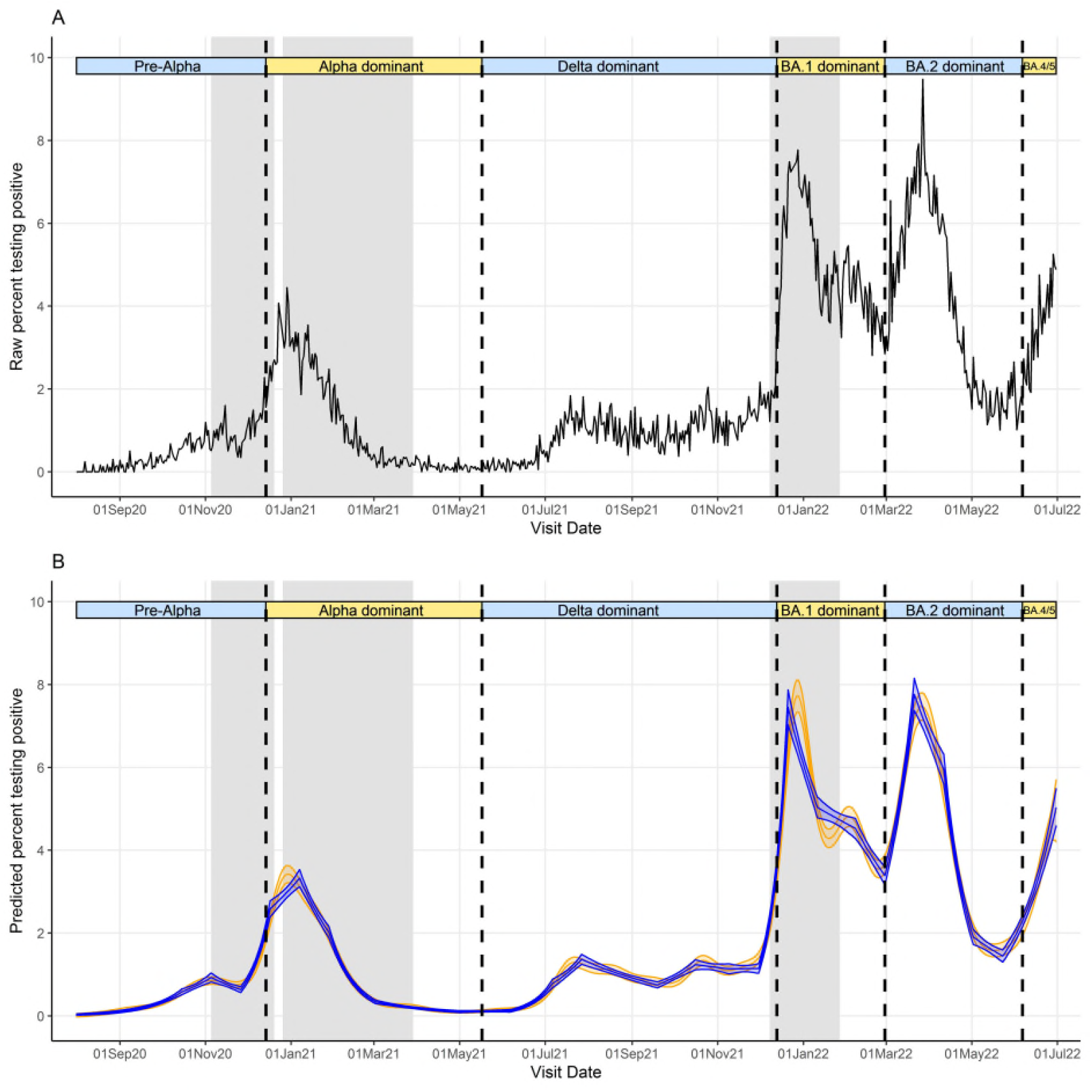
Raw percentage testing positive (A) and predicted percentage of visits testing positive (B) for SARS-CoV-2 from ISR (blue) and GAMs (orange) for London only. Note: Vertical dashed lines indicate periods when new variants became dominant, defined as >50% of positive swabs with cycle threshold (Ct)<30 being S-gene target positive (ORF1ab+N+S, ORF1ab+S, N+S gene positivity) in the Covid-19 Infection Survey for the pre-Alpha period (01 August 2020 - 13 December 2020), the Delta variant (17 May 2021 – 12 December 2021), and the Omicron BA.2 variant (28 February 2022 – 5 June 2022), and >50% Ct<30 S-gene target negative (ORF1ab+N gene positivity) for the Alpha variant (14 December 2020 – 16 May 2021), Omicron BA.1 variant (13 December 2021 – 27 February 2022), and Omicron BA4/BA.5 (6 June 2022 onwards). Gray shaded indicate periods where stay/work from home laws were enforced, although specific restrictions varied across the time series.

### Detection of changes in growth rates using ISR and GAMs

We compared change-points detected across the entire study period by the two methods, first considering emergence of dominant SARS-CoV-2 variants. ISR and GAMs made broadly similar predictions of changing positivity trends across geographical regions over the study period (**Figure 1B, Figure S4**). In London, change-points corresponding to the emergence of Alpha, Delta, BA.1, and BA.2 occurred on 26 November 2020, 6 June 2021, 30 November 2021, and 28 February 2022 using ISR (**Figure 2, Table S3**), and 6 days earlier, 3 days later, 6 and 13 days earlier, respectively, using GAM derivatives. Across all regions, change-points for the three variants were estimated to occur a median 4 days earlier (IQR 0-8) [range 22 days later-26 days earlier] in GAMs versus ISR. 33/48 (69%) of change-points occurred earlier using GAMs. No change-point was detected for Alpha in the East Midlands or Scotland using GAMs, but were detected using ISR.

**Figure 2:**
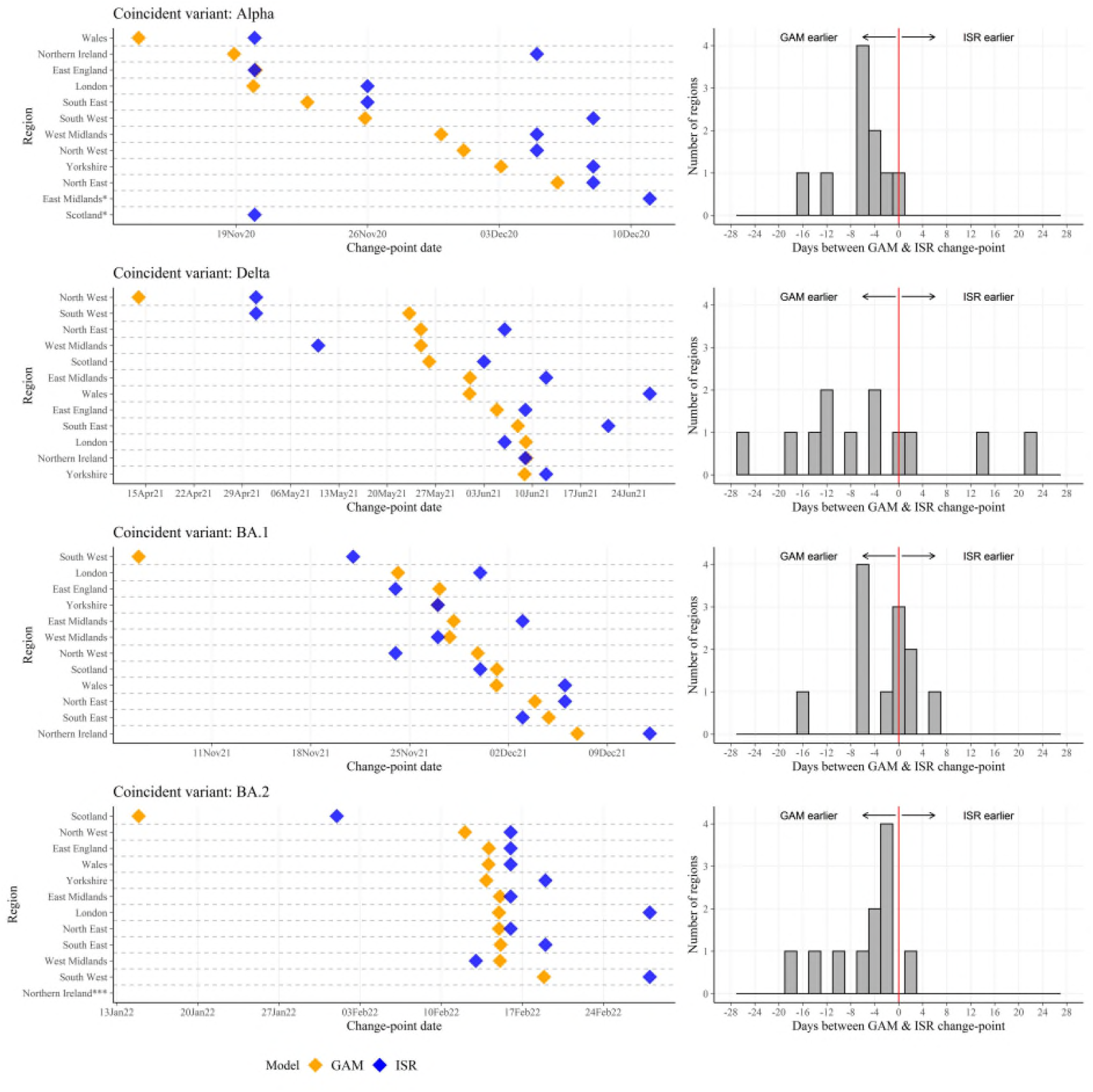
Change-points corresponding to emergence of three key SARS-CoVS-2 variants found by iterative sequential regression (ISR) and second derivatives of generalised additive models (GAM) for each geographical region, run on the full time-series. Note: *No change-point coincident with variant using GAMs; **No change-point coincident with variant using ISR; ***No change-point coincident with variant for GAMs and ISR. Exact dates of change-points are in **Table S3**.

Both methods also identified other change-points aside from trend increases resulting from emergence of these variants, as described for London in **Table 1**. 63% (12/19) of all change-points in London identified in GAMs were identified using ISR within ±7 days. 57% (12/21) of all change-points in London identified by ISR were identified by GAMs within ±7 days. Inconsistent change-points between methods generally reflected small fluctuations when positivity was low.

**Table 1:** All changepoints found by iterative sequential regression (ISR) and second derivatives of generalised additive models (GAM) for London.

| GAM change-point date | ISR change-point date | Description of trend | Days between ISR and GAM change-point |
| --- | --- | --- | --- |
| 26.09.2020 | - | Faster growth | - |
| - | 15.10.2020 | Faster growth | - |
| 02.11.2020 | 05.11.2020 | Increase to decrease | -3 |
| 20.11.2020 | 26.11.2020 | Decrease to increase (rise in Alpha) | -6 |
| 19.12.2020 | 17.12.2020 | Slower growth (slowing down of Alpha) | 2 |
| - | 07.01.2021 | Increase to decrease | - |
| 23.01.2021 | 28.01.2021 | Faster decline | -5 |
| 05.02.2021 | - | Slower decline | - |
| - | 02.03.2021 | Slower decline | - |
| - | 01.05.2021 | Decrease to increase | - |
| 09.06.2021 | 06.06.2021 | Faster growth (rise of Delta) | 3 |
| 12.07.2021 | 06.07.2021 | Slower growth | 6 |
| - | 27.07.2021 | Increase to decrease | - |
| 25.09.2021 | 19.09.2021 | Decrease to increase | 6 |
| 15.10.2021 | 16.10.2021 | Increase to decrease | -1 |
| 01.11.2021 | - | Decrease to increase | - |
| - | 09.11.2021 | Decrease to increase | - |
| 24.11.2021 | 30.11.2021 | Faster growth (rise of BA.1) | -6 |
| 20.12.2021 | 21.12.2021 | Increase to decrease (decline of BA.1) | -1 |
| 06.01.2022 | 11.01.2022 | Slower decline | -5 |
| 29.01.2022 | - | Slower decline | - |
| - | 07.02.2022 | Decrease to increase | - |
| 15.02.2022 | - | Faster growth (rise of BA.2) | - |
| - | 28.02.2022 | Decrease to increase (rise of BA.2) | - |
| 16.03.2022 | 21.03.2022 | Increase to decrease (decline of BA.2) | -5 |
| - | 11.04.2022 | Faster decline | - |
| 19.04.2022 | - | Faster decline | - |
| - | 02.05.2022 | Slower decline | - |
| 26.05.2022 | 23.05.2022 | Decrease to increase (rise of BA.4/BA.5) | 3 |

Note: Change-points were classified as found by both models if within $\pm 7$ days.
\*Negative numbers indicate earlier occurrence of change-points using GAMs, compared with ISR.

### Detection of change-points in ‘near real-time’

While retrospectively detecting change-points could be useful to explore how epidemic growth has varied, ideally change-points would be detected in near real-time to inform measures intended to control growth. Comparing GAMs run on double (16-weeks), triple (24-weeks), or quadruple (32-weeks) an arbitrary but realistic 8-week period of interest showed that 32-weeks data was the minimum that avoided missing over half the change-points in the full time-series (**Table S4, Supplementary Results**).

Running GAMs sequentially adding new data for London every week from 1^st^ October 2020-30th June 2022, 96 change-points were found in the final 8-weeks across all GAMs (**Figure 3**). The majority (64/96: 67%) of change-points were identified by five successive GAMs. Eight (8%) change-points were not identified in any of the five subsequent GAM models, but four of these were identified by ISR. Overall, 77% (74/96) of change-points in the last 8-weeks of successive GAMs were identified by ISR, and 23% (22/96) were never identified by ISR. Results were broadly similar for Northern Ireland (**Supplementary Results**).

**Figure 3:**
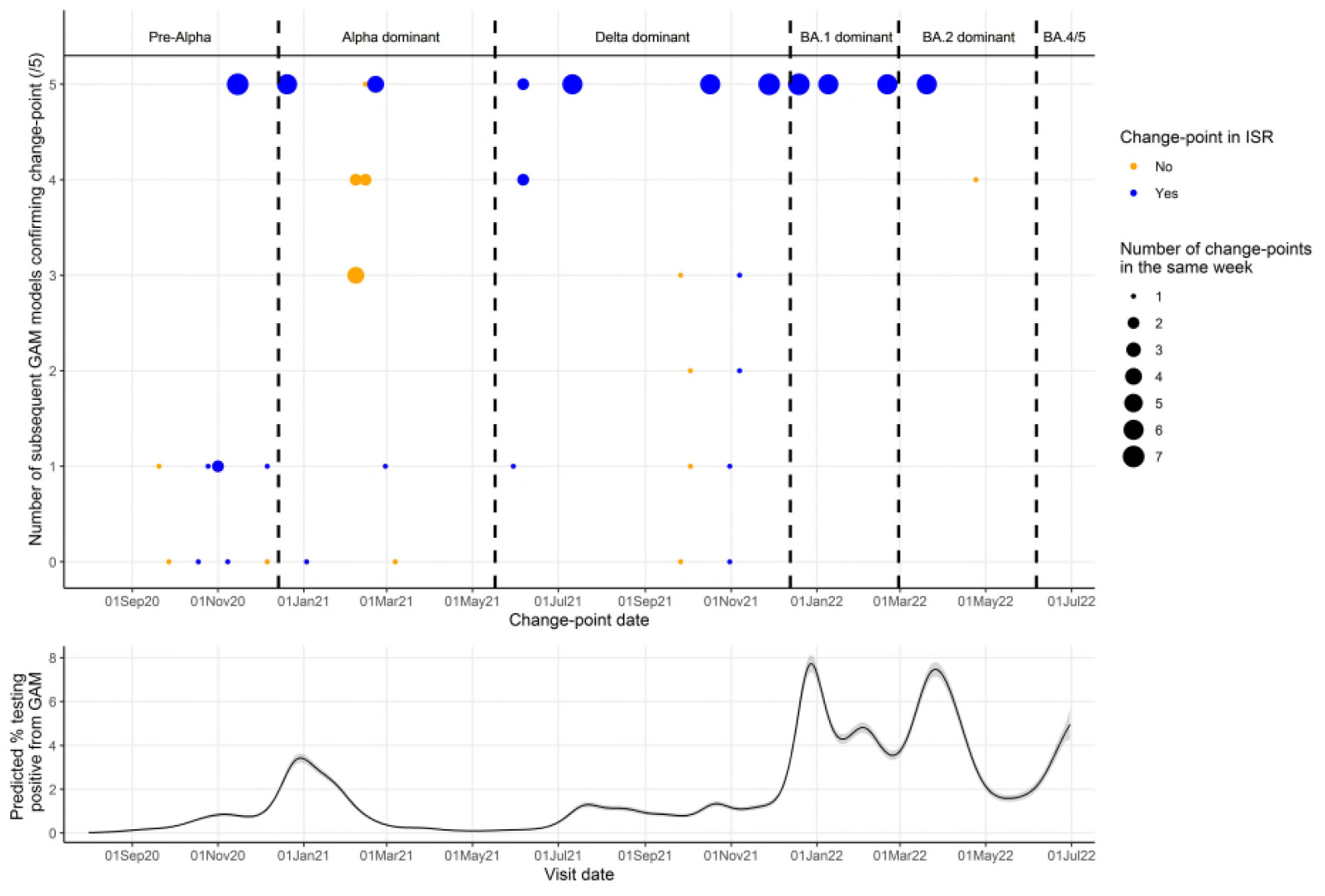
Number of successive GAMs (zero to five), and ISR, finding the same change-points (top panel). Predicted positivity from final GAM for reference (bottom panel). Results are for London only. Note: Change-points in the same week (starting Monday) found in the same number of subsequent models were grouped together (indicated by size of circle). Points are blue if at least one change-point in that week was also found by ISR, and orange is no change-points in that week were found by ISR.

Using the final date of the first successive GAM to estimate when change-points in the full time-series GAM would have been detected, for London, change-points were detected a median 21 (IQR 17-26; range 10-128) days after the change in growth was estimated to occur (**Figure 4; Table S5**). ISR generally fixed change-points into the model (based on lower AIC vs linear trend) 24 days after the change. When identified by both GAMs and ISR, GAMs detected change-points a median 4 (IQR 10 days earlier, 1 day later; range 17 days earlier-35 day later) days earlier. Four change-points identified in the final GAM for London were not identified in any successive GAMs, hence a detection date could not be determined.

**Figure 4:**
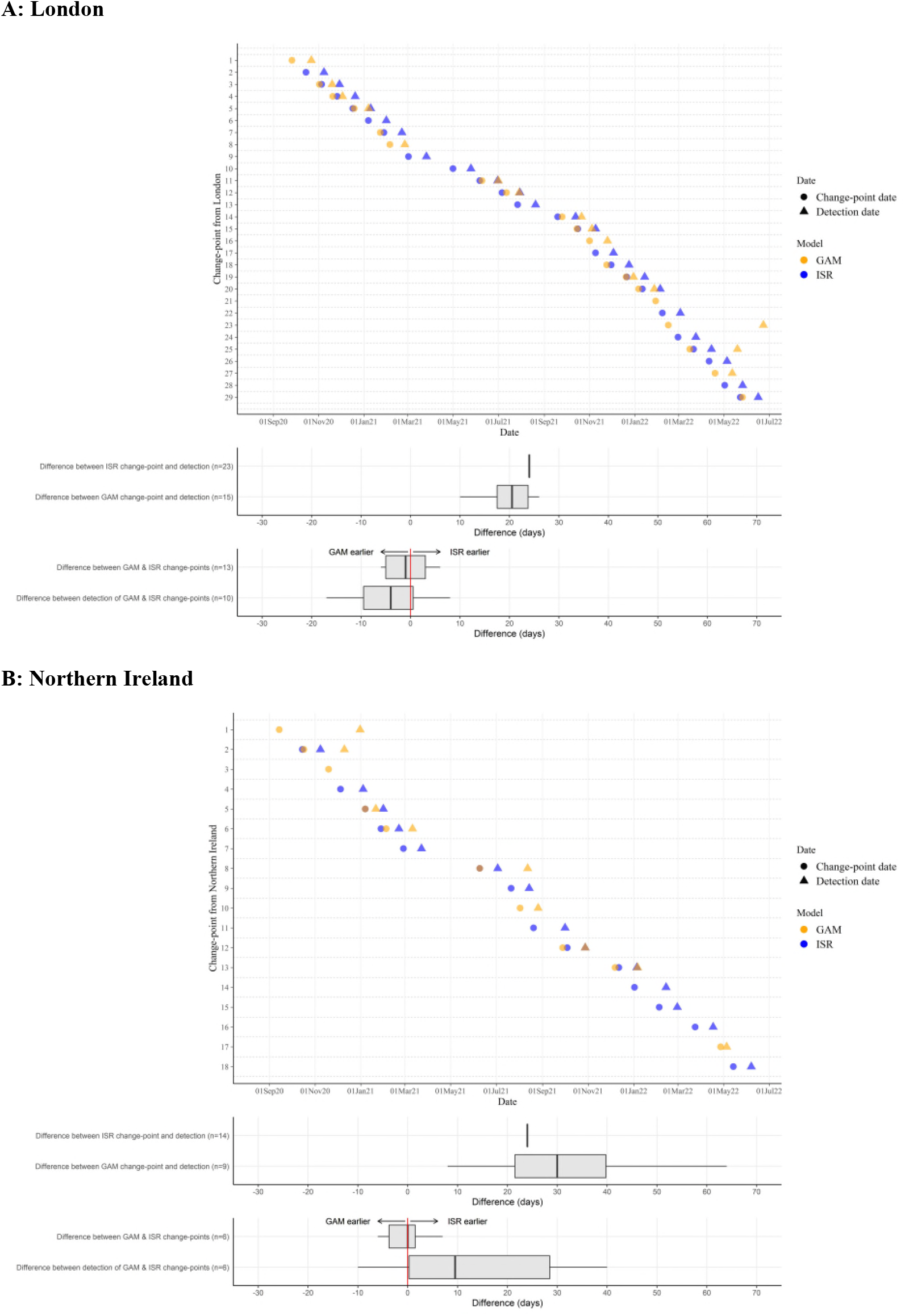
Difference in detection dates between GAMs (orange) and ISR (blue) for London (A) and Northern Ireland (B).

When considering change-points for Northern Ireland (around a fifth of the number of visits in London), while ISR still consistently detected change-points ~24 days after the change occurred, GAMs detected changes median 30 (IQR: 24-54; range: 8-108) days after (**Figure 4; Table S5**). When identified by both ISR and GAMs, in contrast to London, ISR detected change-points a median 10 (IQR: 0, 32) days earlier.

### Incorporating change-points based on the first derivative

Change-points for BA.4/BA.5 were found for all regions using ISR, but were not found using GAMs for 9/12 regions (**Table S6**). Growth rate of BA.4/BA.5 was similar to that of BA.2 declining so, while the second derivative was significantly different from zero, a new change-point was not established. Adding in additional change-points where the first derivative switched signs, all regions found change-points for BA.4/BA.5 using GAMs (**Figure S6**). Further details on additional change-points established by the first derivatives are provided in **Supplementary Results**.

### Estimating change-points in target subgroups

Analogous to “sentinel surveillance”, we assessed whether change-points could be established earlier by modelling population subgroups, here age. In our dataset, as others,^19^ large rises in positivity associated with the emergence of Alpha occurred earlier in those aged 2y-11sy, with steeper increases in positivity in late-August 2021 (Delta) and late-January 2022 (BA.1) compared with older age-groups (**Figure S7, S8**).

Little difference was seen across age groups for GAM change-points associated with Alpha. (**Figure 5**). For Delta, change-points occurred earliest in the overall model and those aged 12sy-49y, and latest in those aged 2y-11sy. Rises in BA.1 occurred 18 days earlier in the youngest age group versus all ages using ISR, and 19 days earlier using GAMs. Rises in BA.2 were found earliest in 2y-11sy ages using GAMs (9 February 2022).

**Figure 5:**
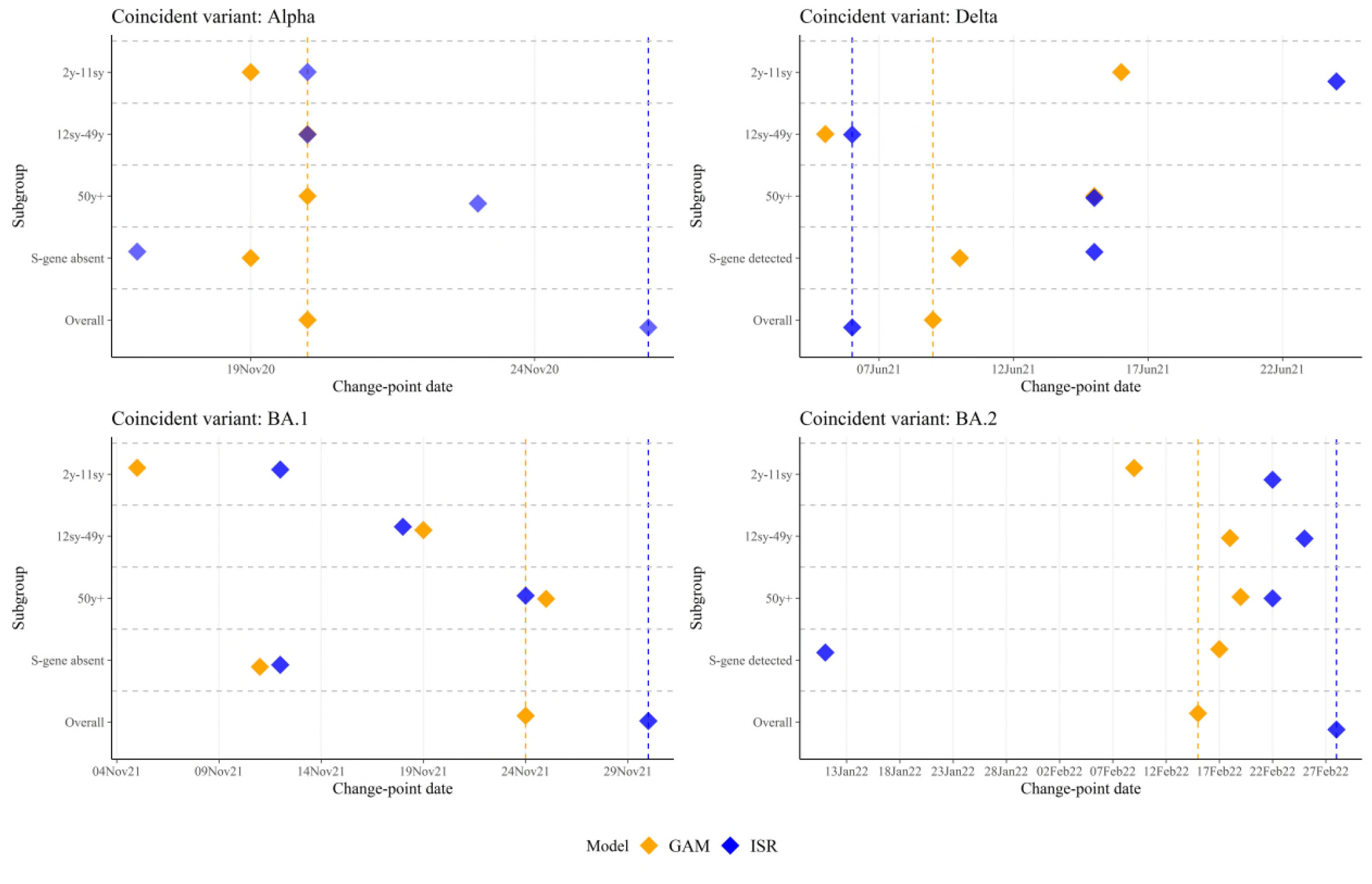
Change-points from GAMs (run on the full time-series; orange) and ISR (blue) run separately by age, separately by S gene detection, and overall in London. Note: Vertical dashed lines show position of change-points in overall GAM and ISR. Change-point dates are provided in **Table S7**.

### Estimating change-points by type of outcome

Analogous to surveillance of different types of infection, e.g. resistant vs susceptible *Staphylococcus aureus*, we considered whether change-points could be established earlier by modelling PCR S-gene positivity as a proxy for SARS-CoV-2 variant. There were distinct differences in trend between SGTF and SGTP positivity over time (**Figure S9, S10**) and GAM and ISR predictions for London closely followed these trends (**Figure S11**).

For London, change-points associated with the emergence of Alpha occurred one and nine days earlier using SGTF versus all positives for GAMs and ISR, respectively (**Figure 5**). Change-points for Delta occurred and were detected nine days later using SGTP versus all positives using ISR, and occurred on one day later using GAMs. Change-points for BA.1 occurred on 11 and 12 November 2021 using GAMs and ISR for SGTF, with all-positive change-points on 24 and 30 November 2021, respectively, a 15-day earlier detection for the ISR change-point. For SGTP, ISR estimated a change-point for BA.2 on 11 January 2022 (detected 4 February 2022) and did not find a change-point for all-positives until 48 days later.

## Discussion

Here, we have compared two methods for detecting changes in growth rates in surveillance data, using SARS-CoV-2 infection as an exemplar. Both methods detected trend increases and decreases associated with Alpha, Delta, BA.1, and BA.2 variants, and other smaller growth rate fluctuations, at similar dates. Considering near real-time analysis, most recent change-points detected using GAMs were found in successive GAMs including five subsequent weeks’ data and using ISR, demonstrating consistency between GAM model runs and methods. However, GAMs needed at least 4 times the duration of data over which there was interest in identifying change-points to provide stable estimates. Change-points were, on average, detected slightly earlier using GAMs versus ISR considering larger geographical regions, but this was not consistent across different sized regions or subgroups. Considering positivity trends separately in different age subgroups allowed earlier detection of BA.1 using data from children alone versus all positives and separately for SGTP of BA.2.

Supporting the use of these methods for sentinel surveillance, we generally found that change-points could be detected earlier through modelling age groups separately, often, but not always, in those aged 2y-11sy. While studies have found no evidence of increased transmission on school premises,^20–23^ the increased person-to-person contact associated with attending school (e.g. public transport, gatherings at school pick-up/drop-off) measurably impact the reproduction number.^24^ Rising SARS-CoV-2 positivity in younger ages may therefore be a useful early warning signal for rises in older age groups, where hospitalisation risk and mortality is higher,^25^ although changes in trends were not consistently identified earlier in younger age groups. Implementing surveillance systems separately by subgroups may therefore be an efficient way to detect changes earlier more generally.

We also found change-points were generally estimated to occur slightly earlier when considering positivity split by S-gene detection. This was particularly useful when BA.2 emerged, as BA.1 declines concealed fast BA.2 growth when combining all positives. More broadly, surveillance of pathogens with different susceptibilities could allow similar shifts in underlying variants to be elucidated.^26^

The methods have much wider applicability to infection surveillance, but SARS-CoV-2 provided an ideal opportunity to test them due to rapid changes in positivity and emergence of variants with different epidemiology. Change-points estimated for Alpha using ISR and GAMs were generally consistent with changing UK public health policy. The first Alpha sequence came from a sample on 20 September 2020, but it was not widely recognized until December after its rapid growth throughout November,^27,28^ with regional lockdowns implemented on 23rd December 2020.^29^ By then, change-points had been detected by ISR in four geographical regions, including London and South East England where Alpha rose earliest and fastest. In contrast, Delta was named a variant of concern on 6 May 2021,^30^ approximately a month earlier than change-points occurred in most regions in our analysis, reflecting its earlier identification through rapid increases in infections in India. Fundamentally ISR and GAMs identify when the epidemiology of an infection changes, which may be independent of or coincident with recognition of a new variant with different transmission potential, virulence, or resistance through either genetic sequencing or changes in epidemiology in other countries. Whilst our methods could be applied to the proportion of genetic sequences which are a specific variant, to date this has generally shown log-linear growth for SARS-CoV-2,^31^ without change-points before a new variant becomes the majority sequence.

Real-time surveillance is mostly concerned with the most recent data, where uncertainty is greatest. Using ISR, the majority of change-points were detected slightly later – a limitation of ISR requiring a minimum number of days between the current and last identified change-point. While GAMs detected some change-points earlier, one limitation is that they also found a small number of change-points during the last 7-days of successive model runs which were not confirmed when adding a further 7-days’ data. The increased flexibility afforded through GAMs may therefore cause false-positives at data boundaries. Requiring at least 7-days of data after a change-point, or confirmation in two successive models, would increase certainty. Some change-points identified by GAMs were significant for short durations e.g. one day, and of small magnitude in the second derivative. Whilst statistically significant, these changes may not be meaningfully different, with policy decisions more likely made on larger changes in growth/decay. Further, using second derivatives of GAMs, change-points for BA.4/BA.5 were mostly not found by the 30th June 2022, but could be established when considering additional change-points based on the first derivative swapping signs. Considering changes in the first derivative may be important to avoid missing change-points moving forward.

In terms of our specific exemplar, demonstrating that relevant change-points can be detected in a randomly sampled community population is useful for SARS-CoV-2 surveillance going forward, as this could trigger targeted testing in different regions and/or age groups to help control spread and identify new variants,^32,33^ ultimately aiming to reduce cases/hospitalisations. The large sample size allowed power to detect change-points, despite relatively low positivity rates, enabling us to compare the two methods. Whilst SARS-CoV-2 is a respiratory virus, the methods are applicable more broadly to different infection surveillance data streams. As regards the methods, estimating derivatives using a Metropolis-Hastings or similar sampler is recommended when modelling outcomes during low prevalence periods.

In summary, ISR and second derivatives of GAMs could potentially detect changes in trend in multiple different types of infections in real-time surveillance, including SARS-CoV-2, but more widely including hospital-acquired infections and antimicrobial resistant pathogens. While both methods gave a generally consistent pattern, some known changes in the epidemiology of SARS-CoV-2 caused by emergence of different variants were identified earlier by GAMs than by ISR and vice-versa, therefore running both methods in parallel would be ideal.

## Data Availability

De-identified study data are available for access by accredited researchers in the ONS Secure Research Service (SRS) for accredited research purposes under part 5, chapter 5 of the Digital Economy Act 2017. For further information about accreditation, contact or visit the SRS website.

## Declaration of interests

DWE declares lecture fees from Gilead outside the submitted work. No other author has a conflict of interest to declare

## Author contributions

ASW, ID, and KBP designed and planned the study. EP, K-DV, DWE, SH, TEAP, PCM, NS, KBP, and ASW contributed to the statistical analysis. All authors contributed to interpretation of data and revised the report. All authors approved the final version of the report and agree to be accountable for all aspects of the work.

### Funding

This study is funded by the Department of Health and Social Care and the UK Health Security Agency with in-kind support from the Welsh Government, the Department of Health on behalf of the Northern Ireland Government and the Scottish Government. EP, K-DV, KBP, ASW, TEAP, NS, DWE are supported by the National Institute for Health Research Health Protection Research Unit (NIHR HPRU) in Healthcare Associated Infections and Antimicrobial Resistance at the University of Oxford in partnership with Public Health England (PHE) (NIHR200915). ASW and TEAP are also supported by the NIHR Oxford Biomedical Research Centre. KBP is also supported by the Huo Family Foundation. ASW is also supported by core support from the Medical Research Council UK to the MRC Clinical Trials Unit [MC_UU_12023/22] and is an NIHR Senior Investigator. PCM is funded by Wellcome (intermediate fellowship, grant ref 110110/Z/15/Z) and holds an NIHR Oxford BRC Senior Fellowship award. DWE is supported by a Robertson Fellowship and an NIHR Oxford BRC Senior Fellowship. NS is an Oxford Martin Fellow and an NIHR Oxford BRC Senior Fellow. The views expressed are those of the authors and not necessarily those of the National Health Service, NIHR, Department of Health, or PHE. The funder/sponsor did not have any role in the design and conduct of the study; collection, management, analysis, and interpretation of the data; preparation, review, or approval of the manuscript; and decision to submit the manuscript for publication. All authors had full access to all data analysis outputs (reports and tables) and take responsibility for their integrity and accuracy. For the purpose of Open Access, the author has applied a CC BY public copyright licence to any Author Accepted Manuscript version arising from this submission.

## Notes

### Author Declarations

The study received ethical approval from the South Central Berkshire B Research Ethics Committee (20/SC/0195).

