## Supplementary Material for "Detecting changes in population trends in infection surveillance using community SARS-CoV-2 prevalence as an exemplar"

### Supplementary Methods

#### ISR algorithm

ISR initially fitted a linear trend on the log scale up to 1st September 2020 allowing no change-points to be found in this period. The model then considered the subsequent *n* days of the time-series, and fit two models: one extending the current trend (one-trend), the second allowing a change of trend (two-trend). If the two-trend reduced the AIC by at least 6·635, the change-point was permanently fixed in the model, otherwise the one-trend model was chosen. A minimum length of time between change-points (interval length) had to be chosen, as well as a minimum length of time between a change-point and its detection time (minimum distance). If the one-trend model was selected, the endpoint was moved forward one day and models with one extra change-point at all possible positions were fitted, with the one with the smallest AIC being identified (this could again be the model with no change-points), as well as all the models with an AIC within 6·635. The algorithm repeated this process until the end of the time-series.

#### Second derivative estimation by simulation

Derivatives were estimated for the smooth function using posterior simulation on the absolute scale. If positivity was relatively common throughout the entire period, coefficients from the GAM would approximately follow a multivariate normal distribution with mean vector and covariance matrix specified by the model estimates of the coefficients and their covariances, respectively.^13^ Posterior simulation involves taking random draws from this distribution, whereby each draw represents a new trend that is consistent with the fitted model while also incorporating uncertainty in the estimated trend. However, this Gaussian approximation will be poor in periods where data consists of mostly zeros due to low positivity, as observed for some periods in our exemplar. To overcome this problem, we used a simple Metropolis-Hastings sampler to generate samples from the posterior distribution of the fitted model (as implemented in the gam.mh function from the *mgcv* R package).^16,17^ This approach alternates fixed proposals – based on the typical Gaussian approximation to the posterior – with random walk proposals, based on a shrunken version of the approximate posterior covariance matrix. The random walk component ensures that the chain does not get stuck in regions for which the Gaussian proposal density is much lower than the posterior density.^16^

First, 2000 curves were simulated from the fitted GAMs using a Metropolis-Hastings algorithm. First and second derivatives on the absolute scale were then estimated for each simulation using backwards finite differences, allowing estimation on the final day of data (not possible with forward of central differences). The median, 2·5th, and 97·5th percentiles were estimated across the simulations, obtaining an average with credible intervals

### Supplementary Results

#### Detection of change-points in near real-time: running GAMs on shorter time periods

In real-time, often most interest is in change-points at the end of a time-series, for example, the final 8-weeks. Rather than running GAMs from the start of the time series (1st August 2020) each time we wanted to find new change-points at the end of the time-series, to improve computational efficiency we assessed whether the same change-points were estimated if GAMs were only run on double (16-weeks), triple (24-weeks), or quadruple (32-weeks) the period of interest. The shorter time-frames of 16-weeks and 24-weeks missed over half the change-points in the full time-series in both cases so were not considered further (**Table S4**).

In contrast, the 32-week model found the majority of change-points in the full time-series (8/10). In the 32-week model, one change-point on the 21 February 2021 was not identified by the full model in the model ending 18 March 2021. This change in the second derivative was only significant for two days, thus may not be a meaningful change. The second derivative became significant again on the 24 February 2021 for nine days, matching the change-point in the full model on the 23 February 2021. With models ending on 23 December 2021 and 17 February 2022, the 32-week model missed two change-points identified in the full time-series (4 Nov 2021, 2 Feb 2022). While these were not change-points indicating substantial growth/decay of variants, they were both identified by ISR (9 Nov 2021, 7 Feb 2022; **Table 1**). Thus, while 32-weeks appeared to identify the majority of change-points, if the capacity is available to run models on the full time-series, statistical power will likely be increased.

#### Detection of change-points in ‘near real-time’ for Northern Ireland

Comparing to Northern Ireland (the smallest region in our dataset), 52 change-points were found in the final 8-weeks across all GAMs (**Figure S5**) The majority (31/52: 60%) of these change-points were identified by five successive GAMs. One change-point was not identified in any of the five subsequent GAM models, but was identified by ISR. Overall, 52% (27/52) of change-points in the last 8-weeks of successive GAMs were identified by ISR, and 48% (25/52) were never identified by ISR.

#### Incorporating additional change-points based on the first derivative

Adding in additional change-points where the first derivative switched signs during a period of significance in the second derivative, an additional 78 change-points were established across the full time-series for all regions (199 change-points based on the second derivative only). The largest number of additional change-points occurred in South East England, with 10 additional change-points above the 20 original change-points established using the second derivative only (**Table S6**). The majority of the additional change-points occurred in January and February 2022, concurrent with the rise and fall of BA.1 (**Figure S6**).

### Supplementary Figures

#### Figure S1: Comparison of GAMs with region included at an interaction with time (red) and as separate models for each region (blue)

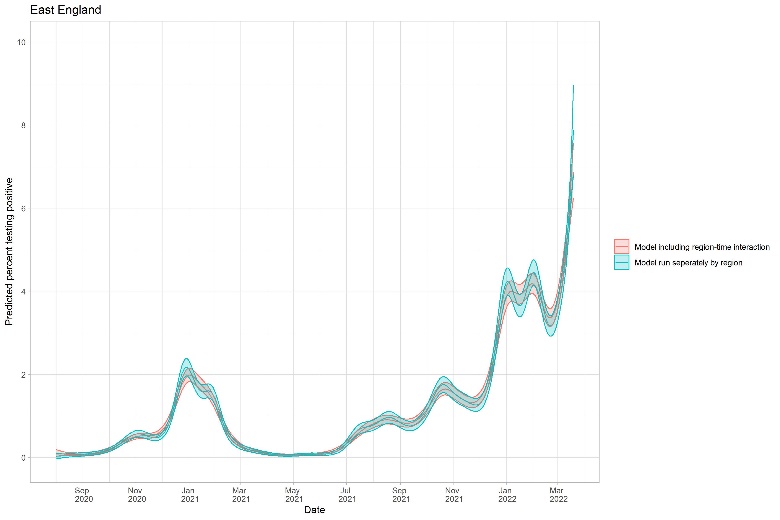

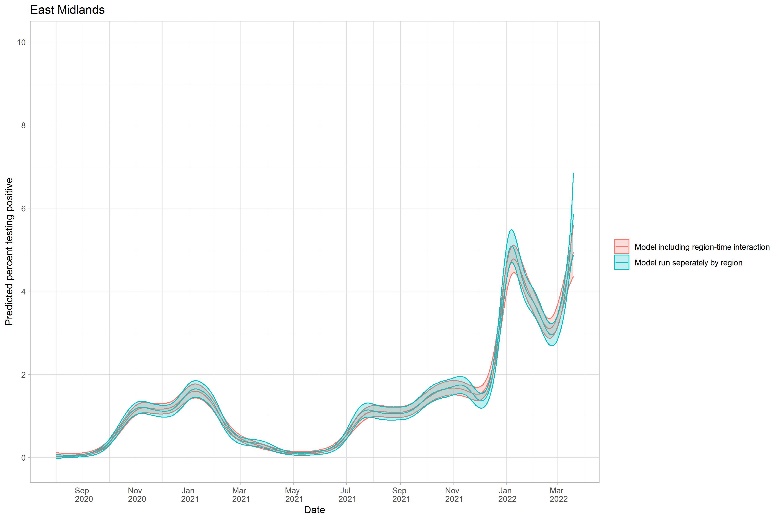

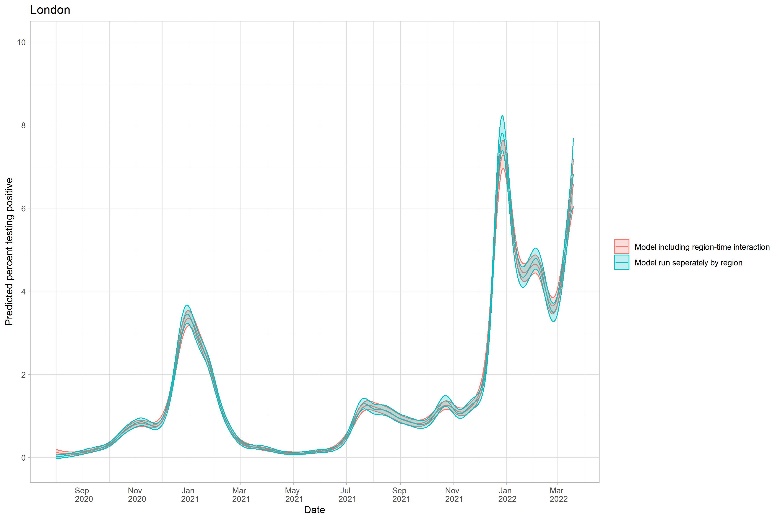

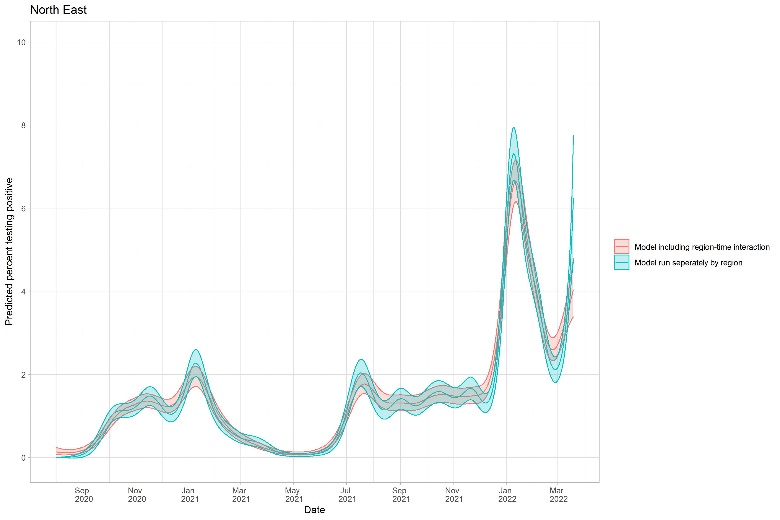

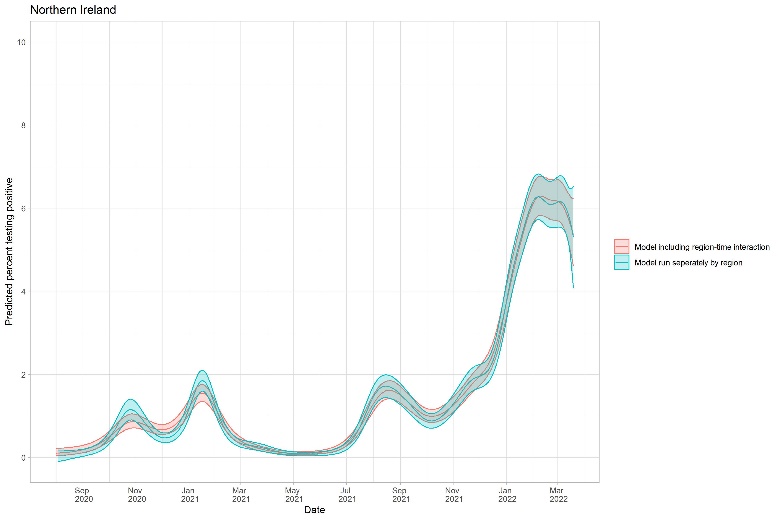

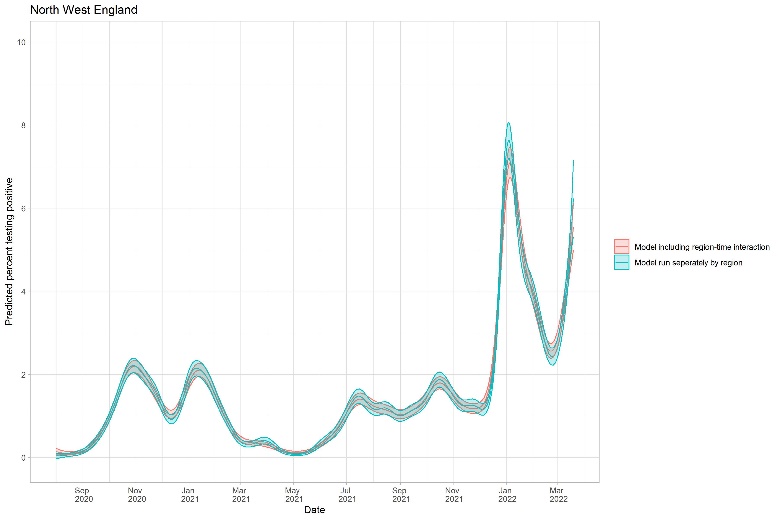

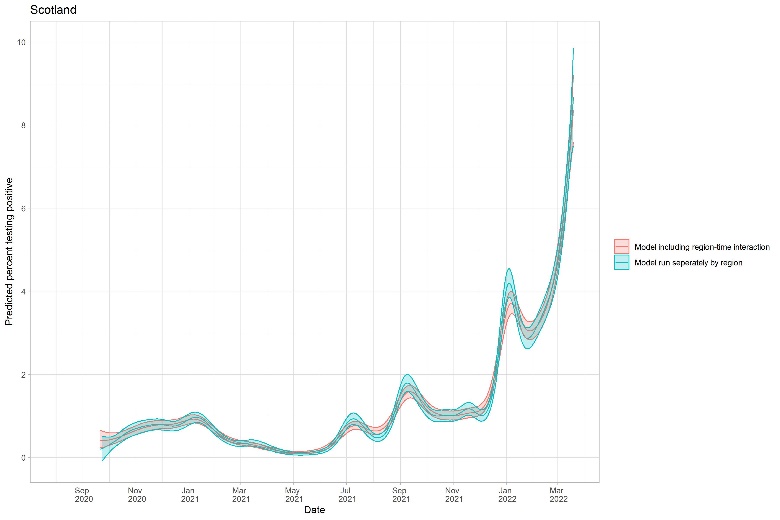

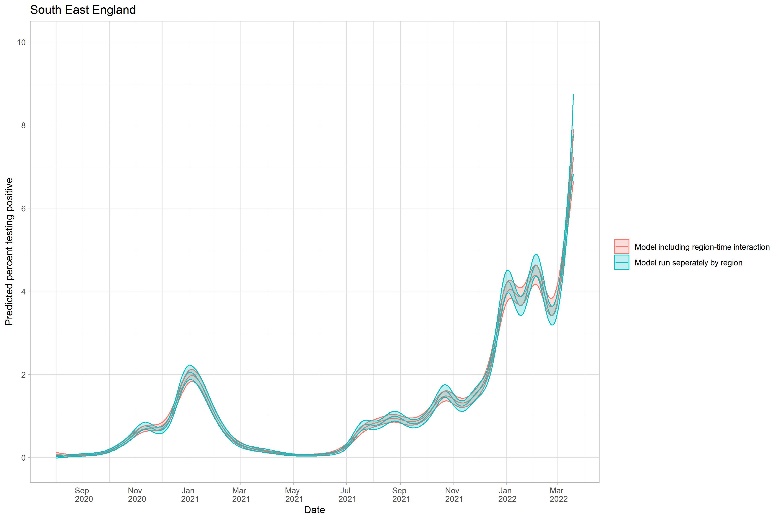

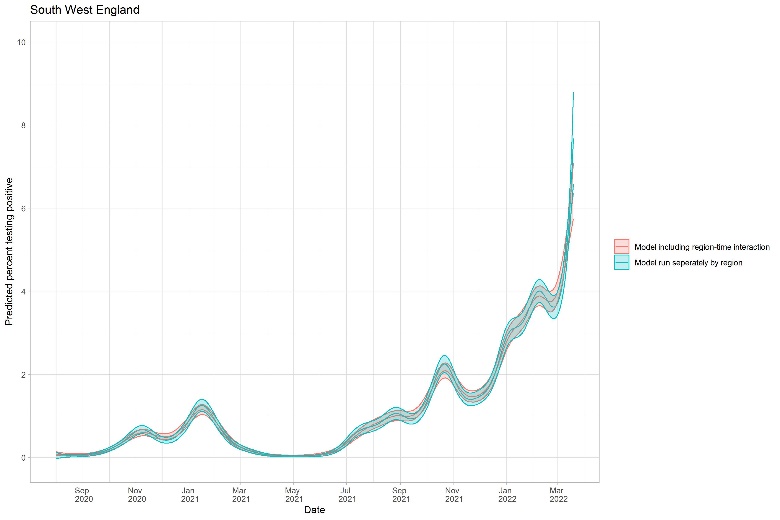

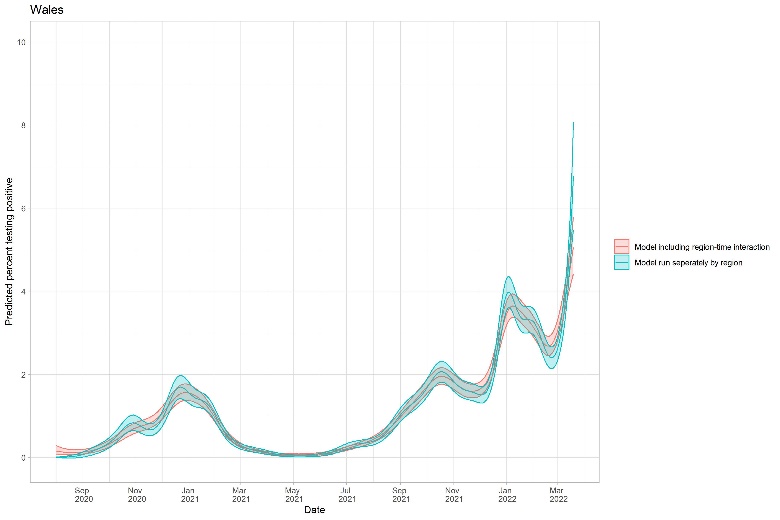

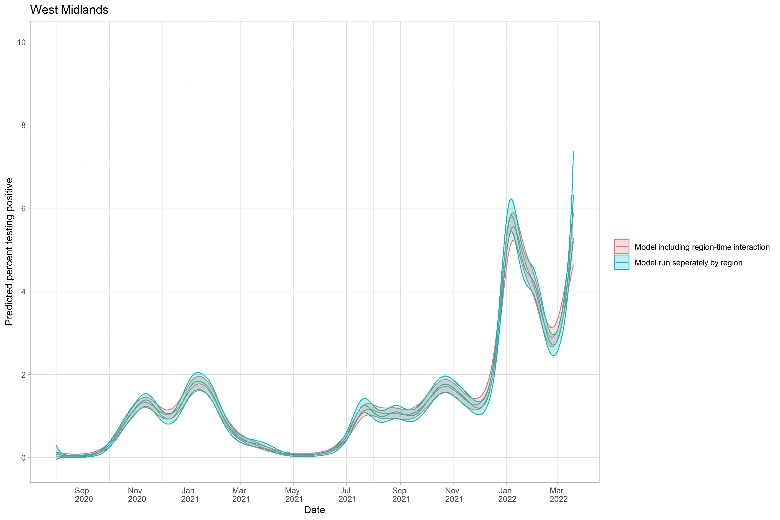

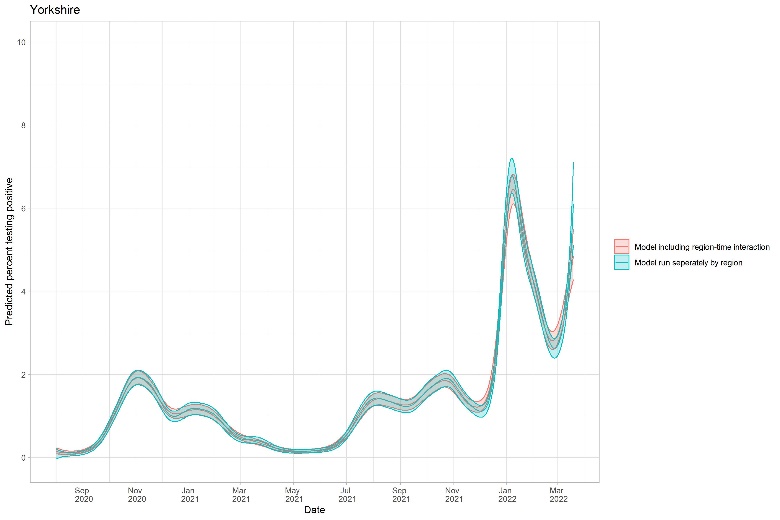

Note: The run-time for the model including the interaction between region-time was 84·6 hours, versus approximately 4 hours when running all regions separately, before derivatives were estimated.

#### Figure S2: Difference in predicted percentage testing positive from GAMs with varying numbers of basis functions (k) of 25, 50, 75, 100, for London only

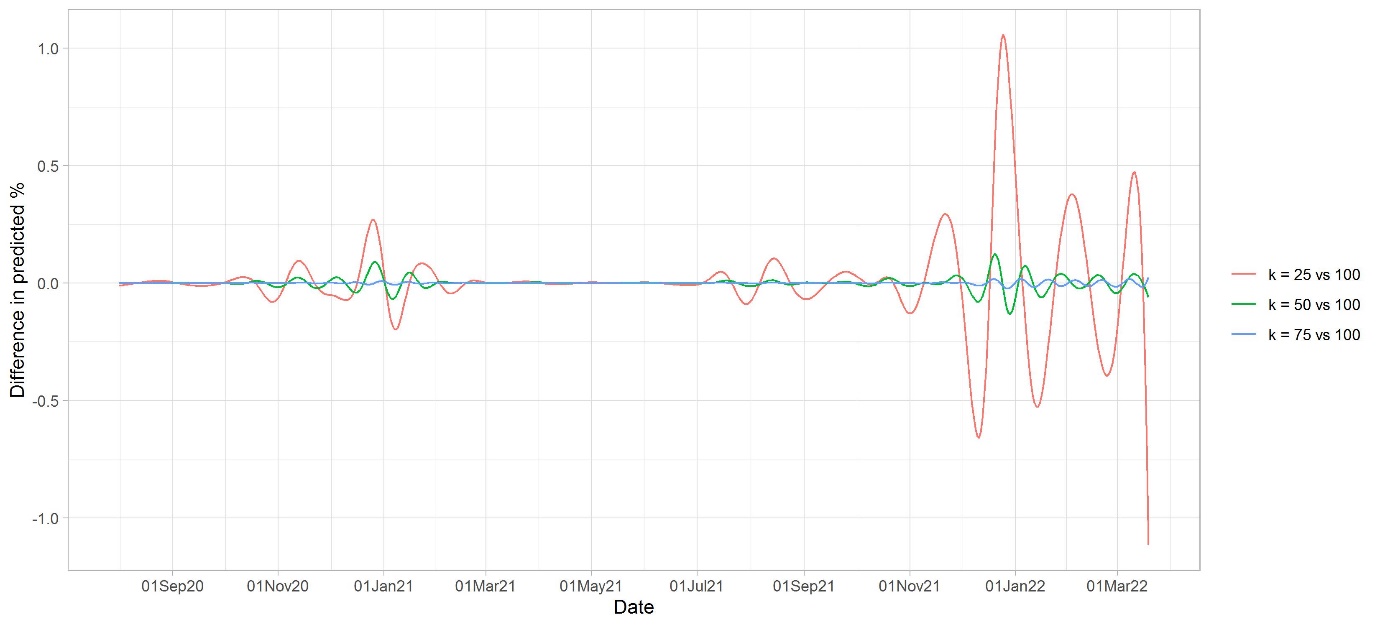

Note: The median (IQR) [range] of differences between GAMs with k = 25, 50, 75 vs k = 100 were -0·0005 (-0·08, 0·01) [-1·09, 1·79], 0·0002 (-0·007, 0·010) [-0·1287, 0·2087], and 0·000007 (-0·0015, 0·0017) [-0·036, 0·032], respectively. The effective degrees of freedom (EDF) were 23·4, 39·5, 44·6, and 45·6 for k = 25, 50, 75, and 100, respectively.

#### Figure S3: Raw daily percentage of visits with a SARS-CoV-2 positive test over the study period overall (A) and split by region (B)

**A: Raw percentage testing positive for all regions combined**

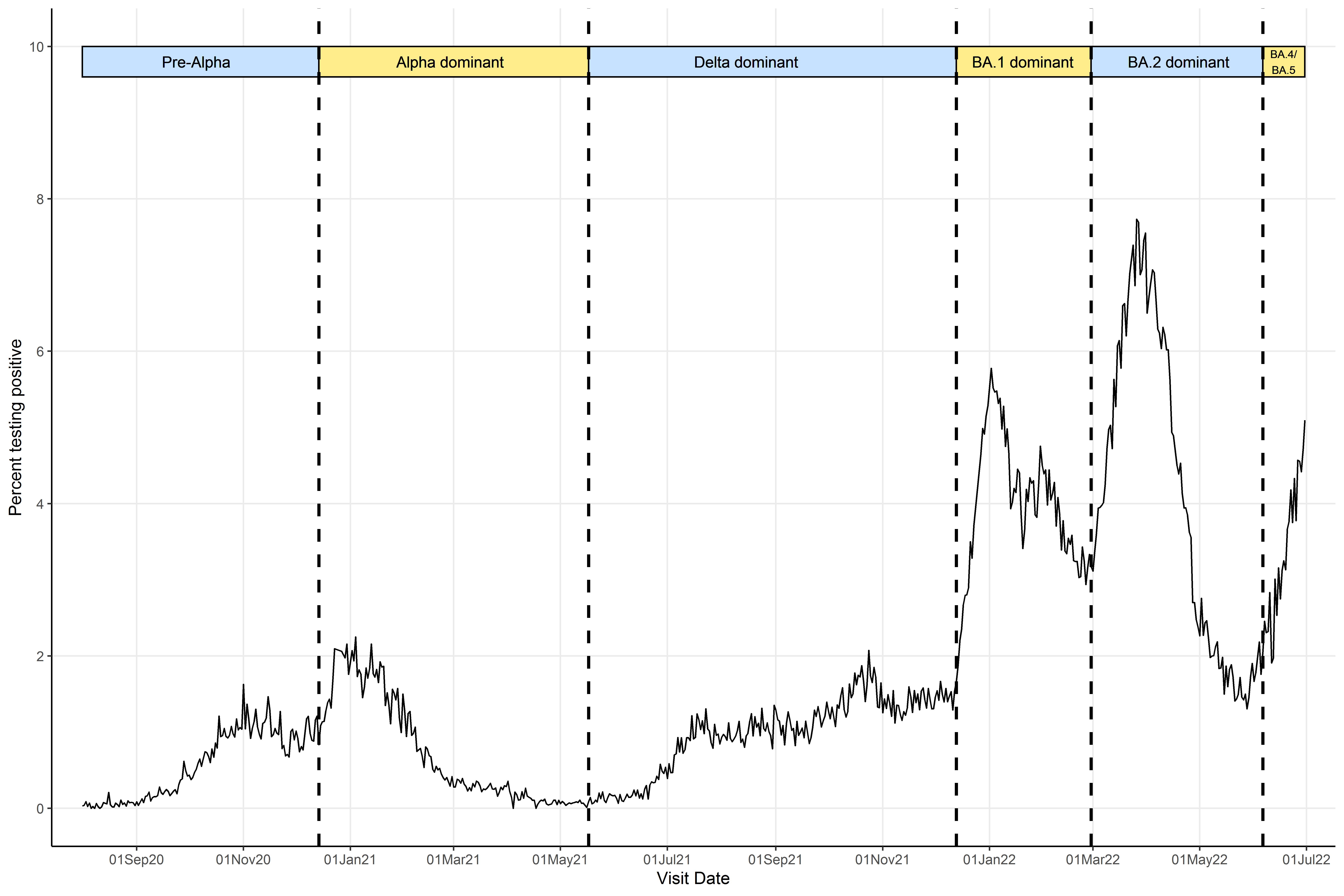

**B: Raw percentage testing positive for each region**

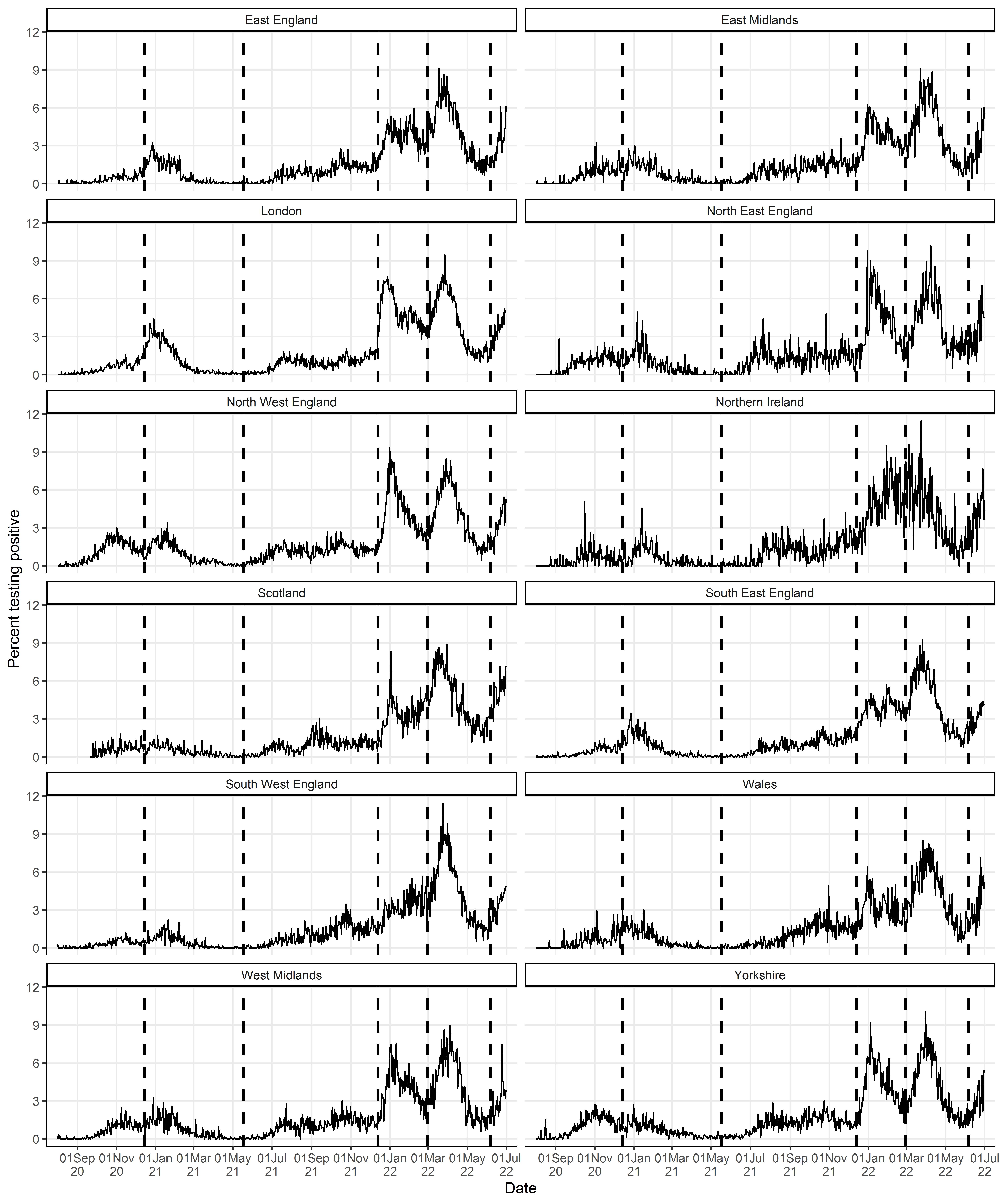

Note: Vertical dashed lines indicate periods when new variants became dominant, defined as >50% of positive swabs with cycle threshold (Ct)<30 being S-gene target positive (ORF1ab+N+S, ORF1ab+S, N+S gene positivity) in the Covid-19 Infection Survey for the pre-Alpha period (01 August 2020 - 13 December 2020), the Delta variant (17 May 2021 – 12 December 2021), and the Omicron BA.2 variant (28 February 2022 – 5 June 2022), and >50% Ct<30 S-gene target negative (ORF1ab+N gene positivity) for the Alpha variant (14 December 2020 – 16 May 2021), Omicron BA.1 variant (13 December 2021 – 27 February 2022), and Omicron BA4/BA.5 (6 June 2022 onwards) .

#### Figure S4: Predicted percentage of visits testing positive for SARS-CoV-2 from ISR (blue) and GAMs (orange) for all regions

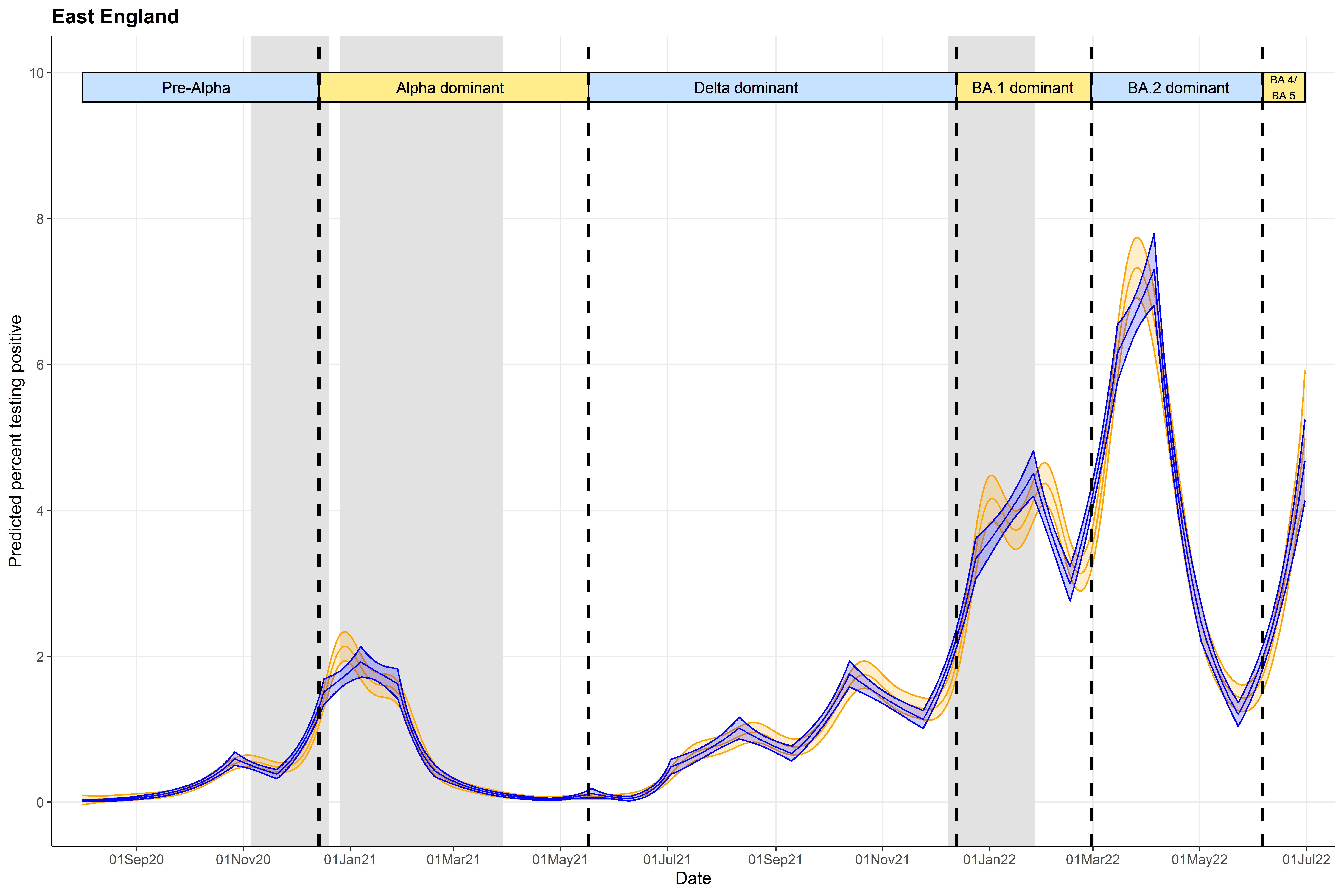

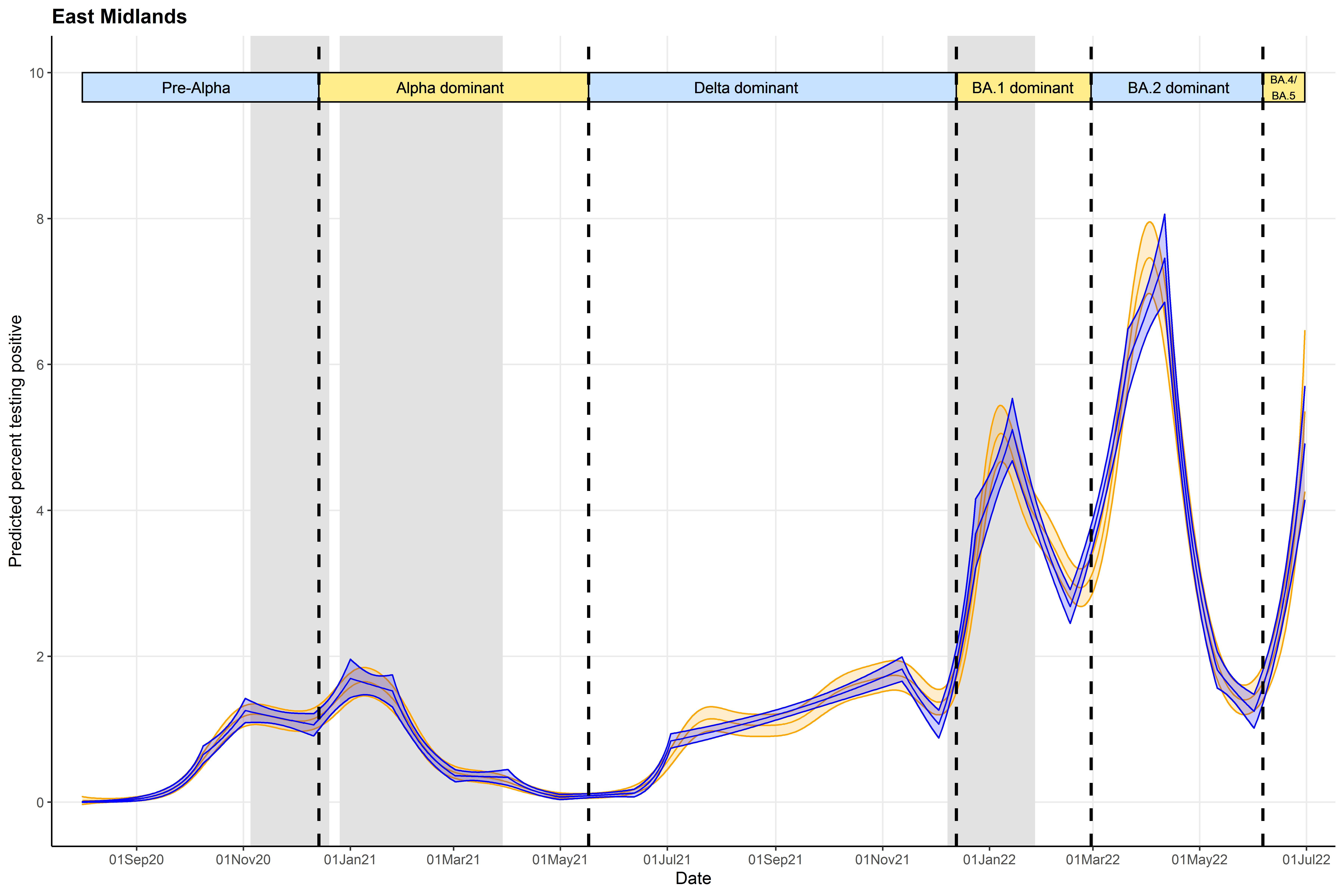

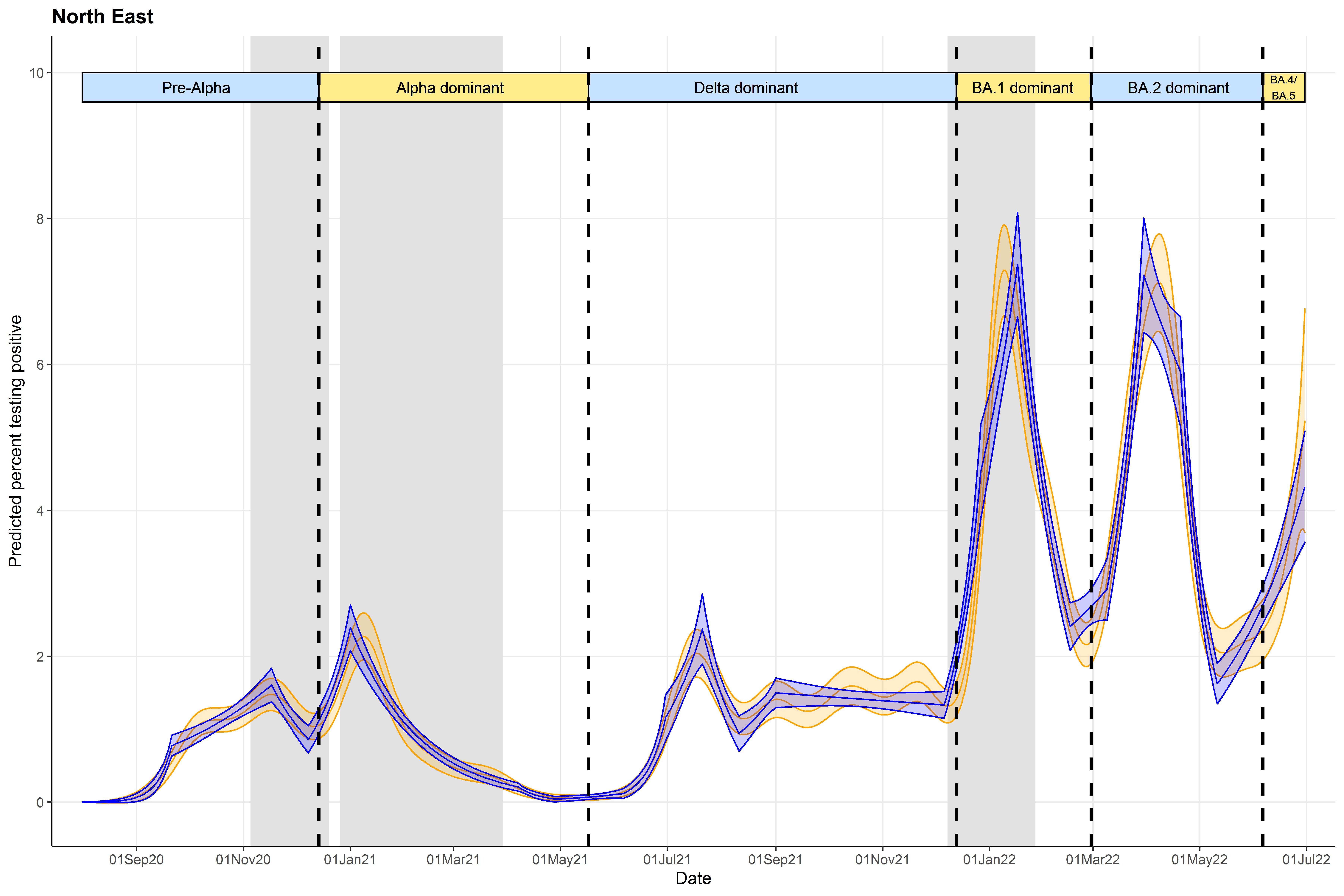

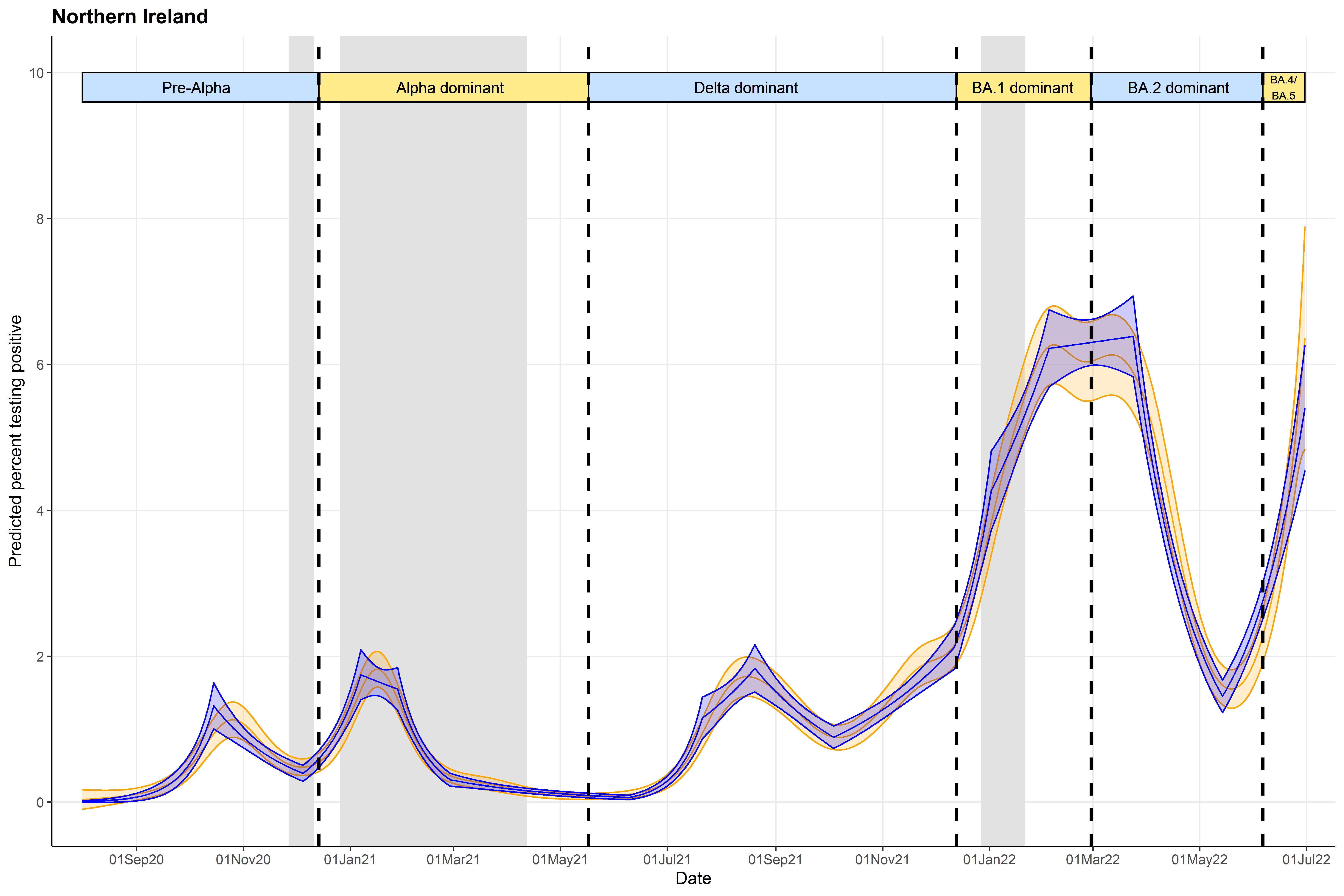

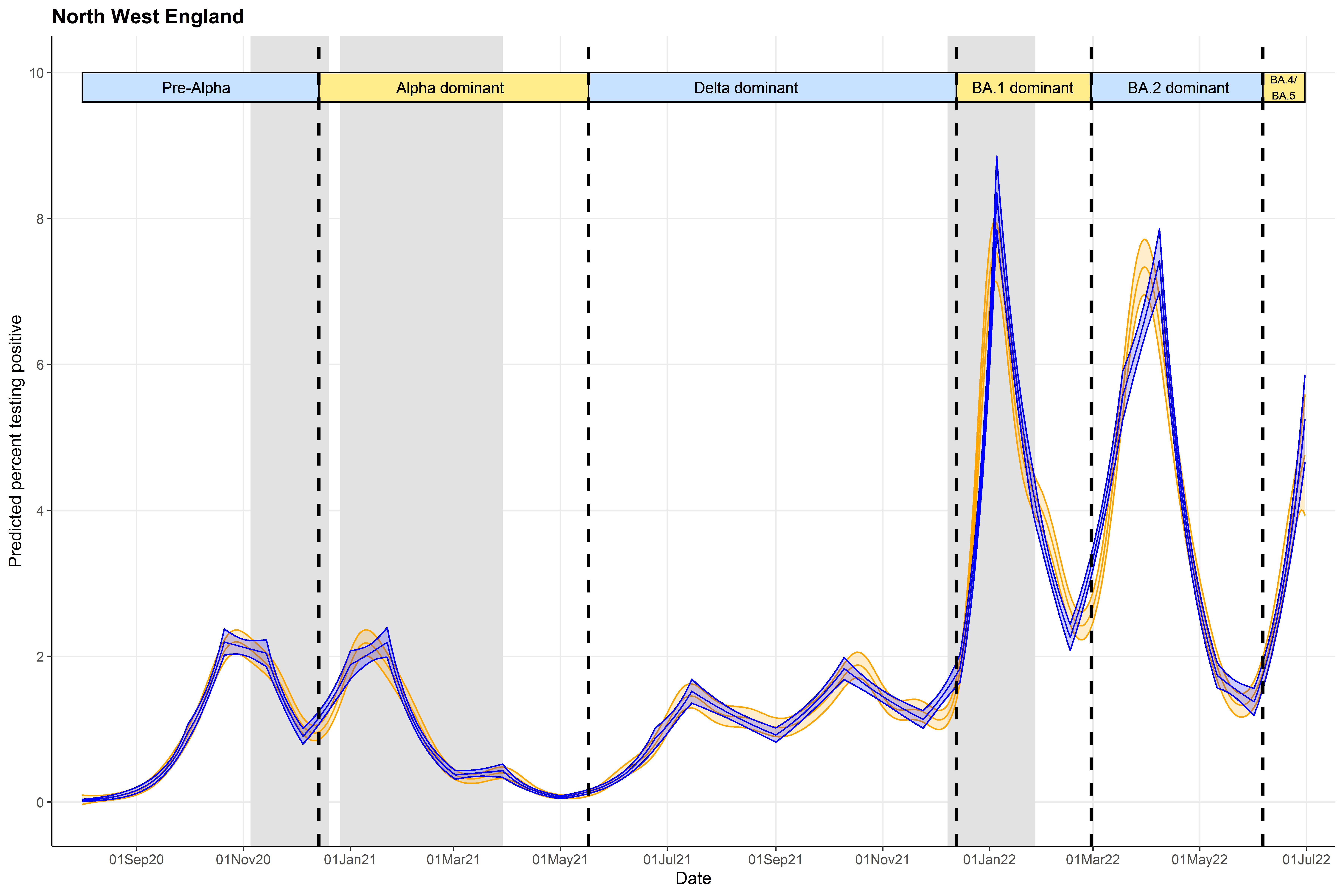

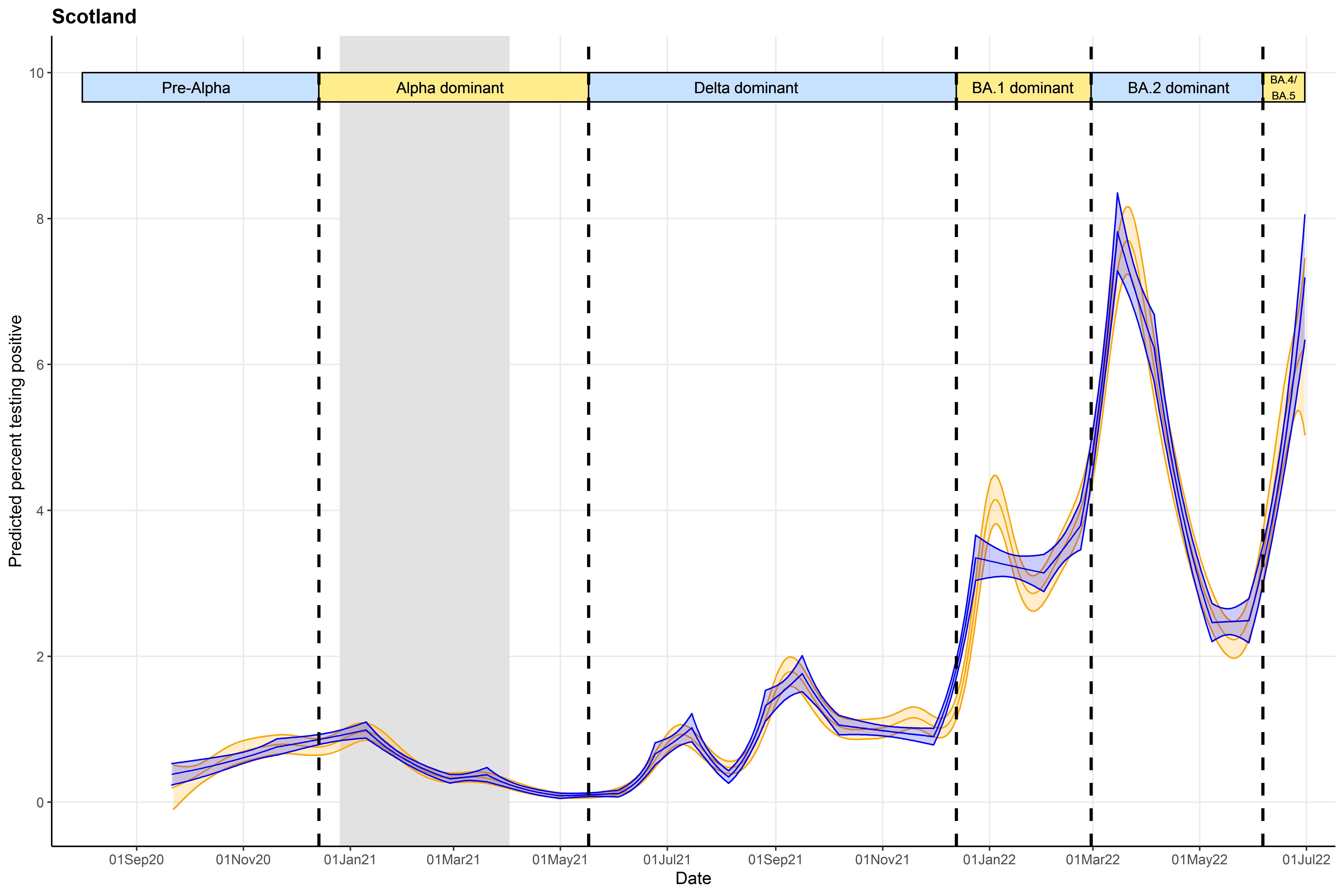

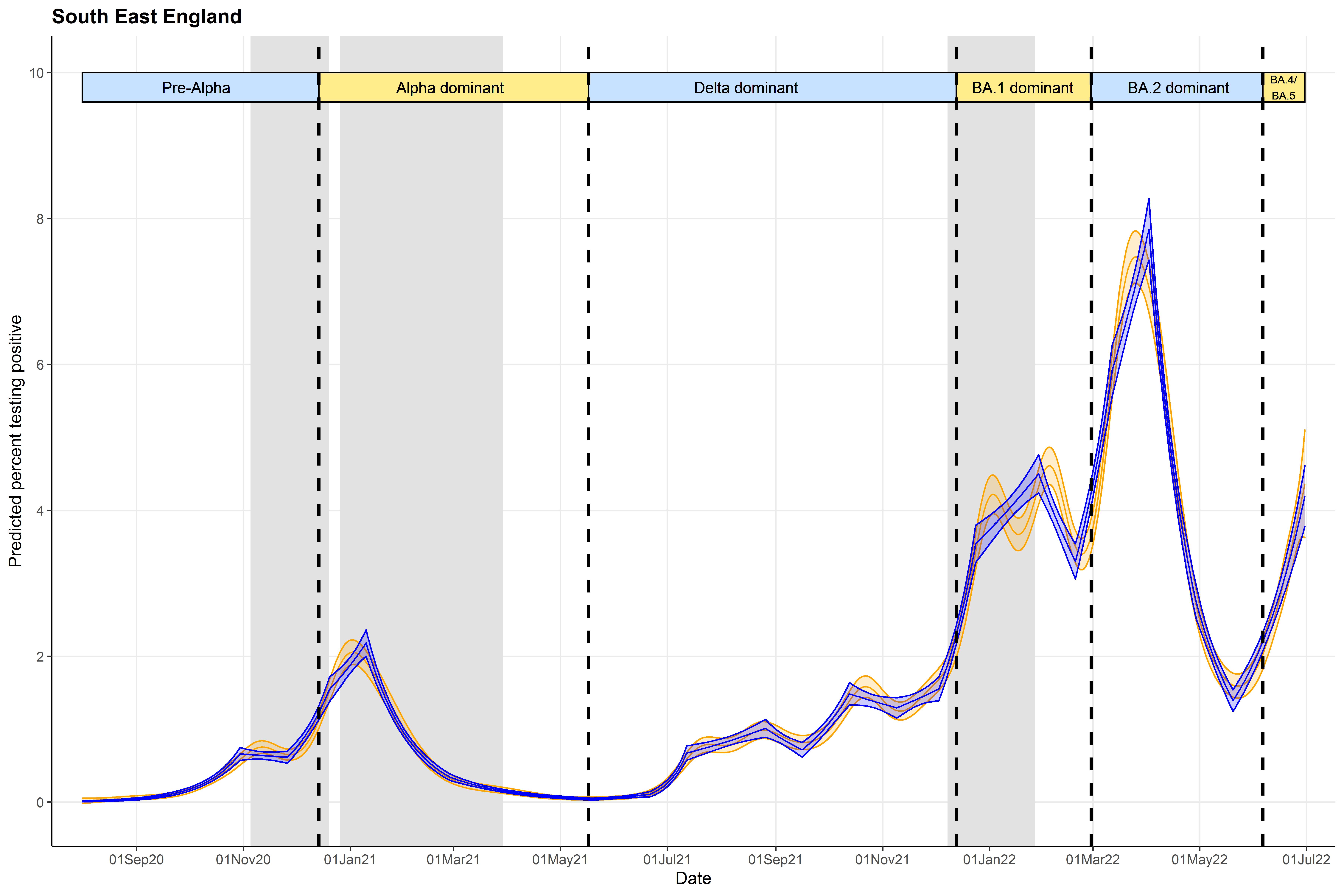

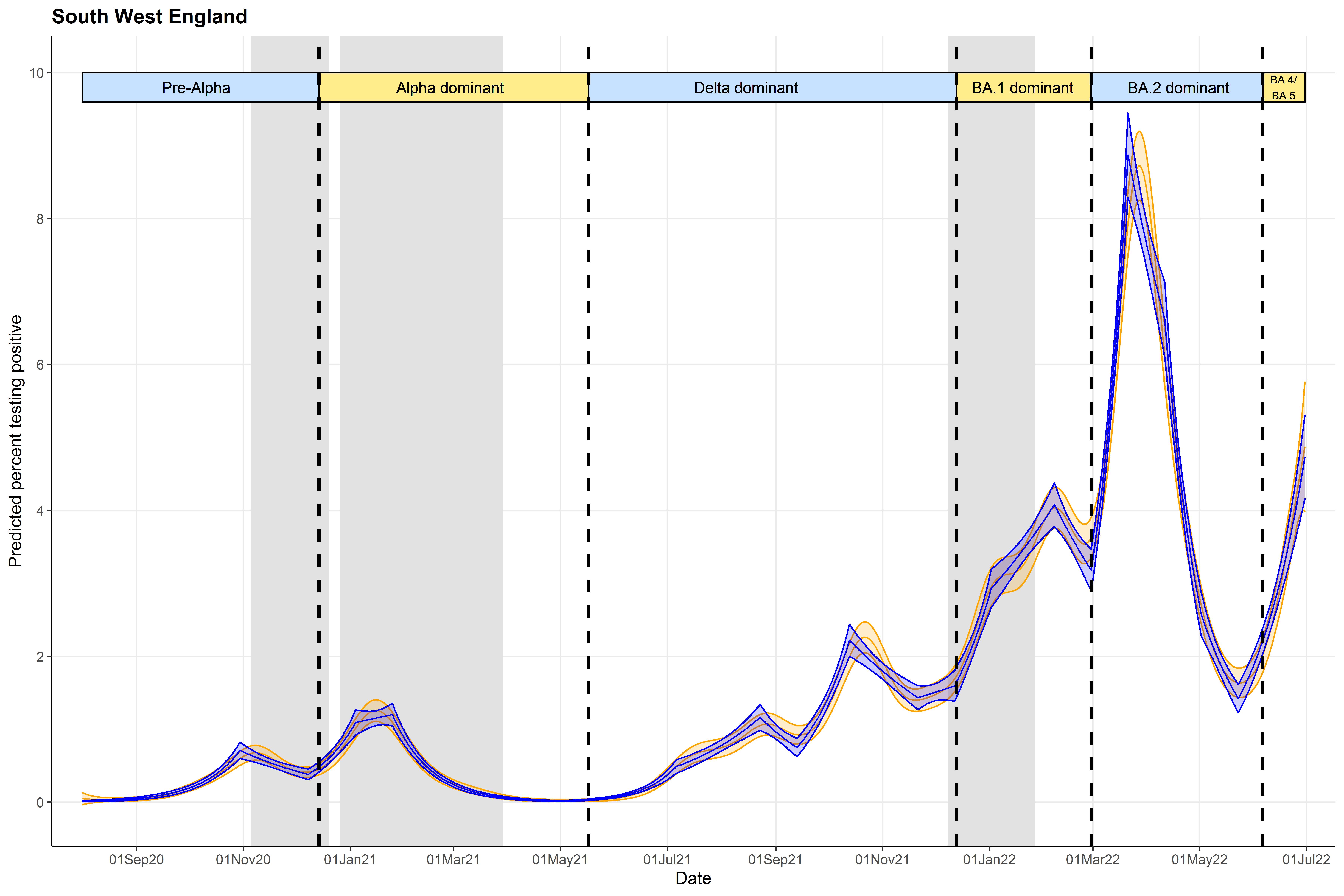

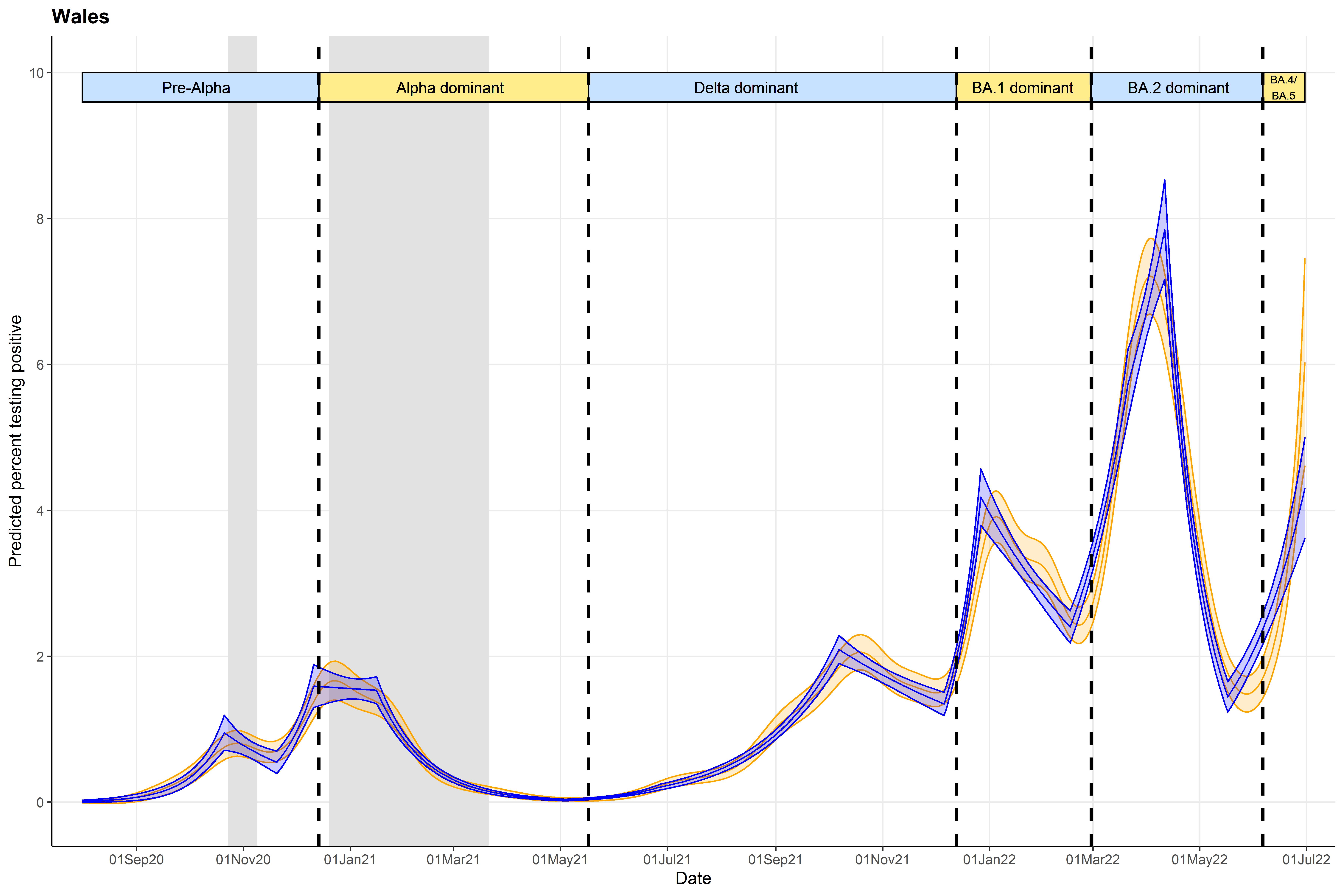

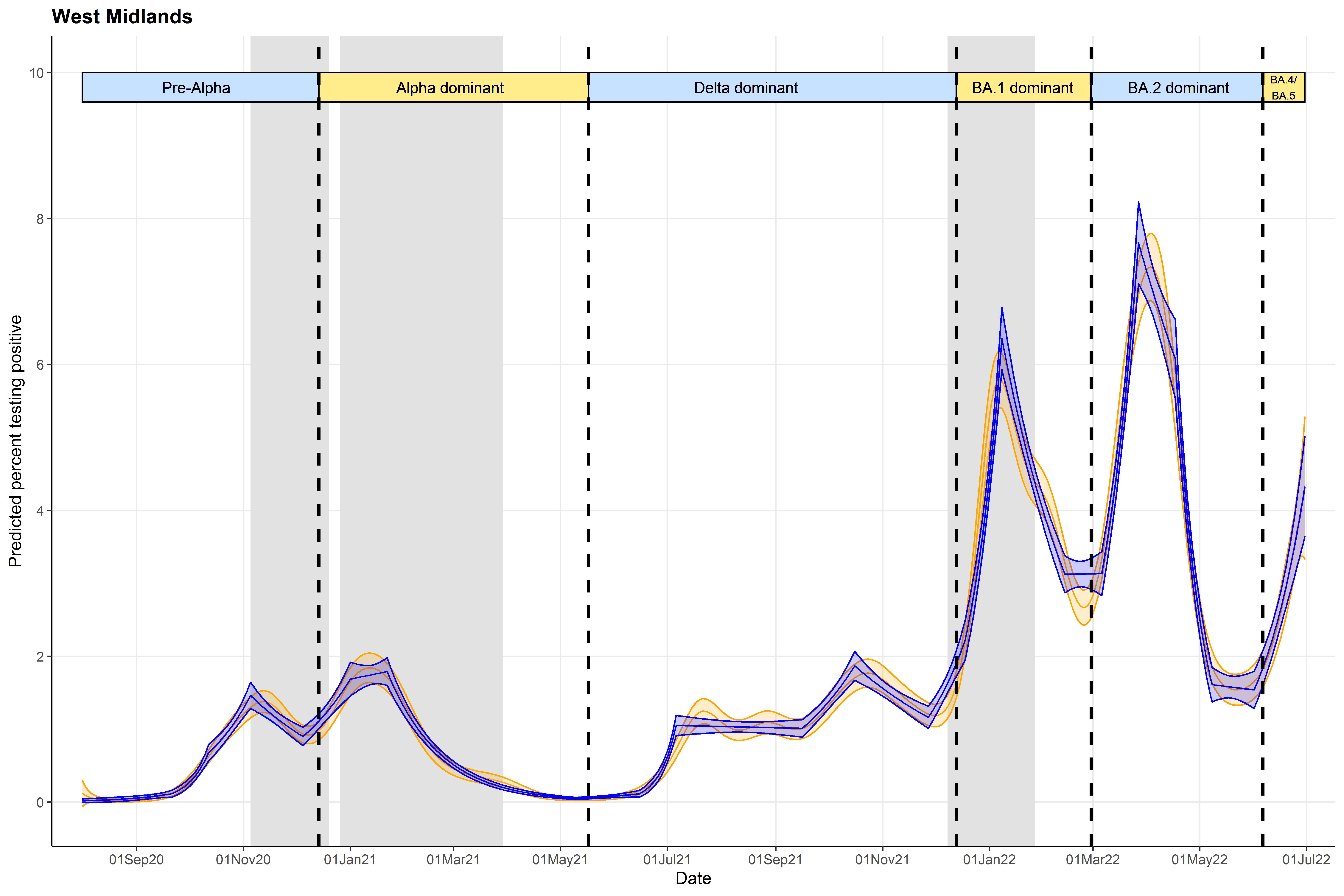

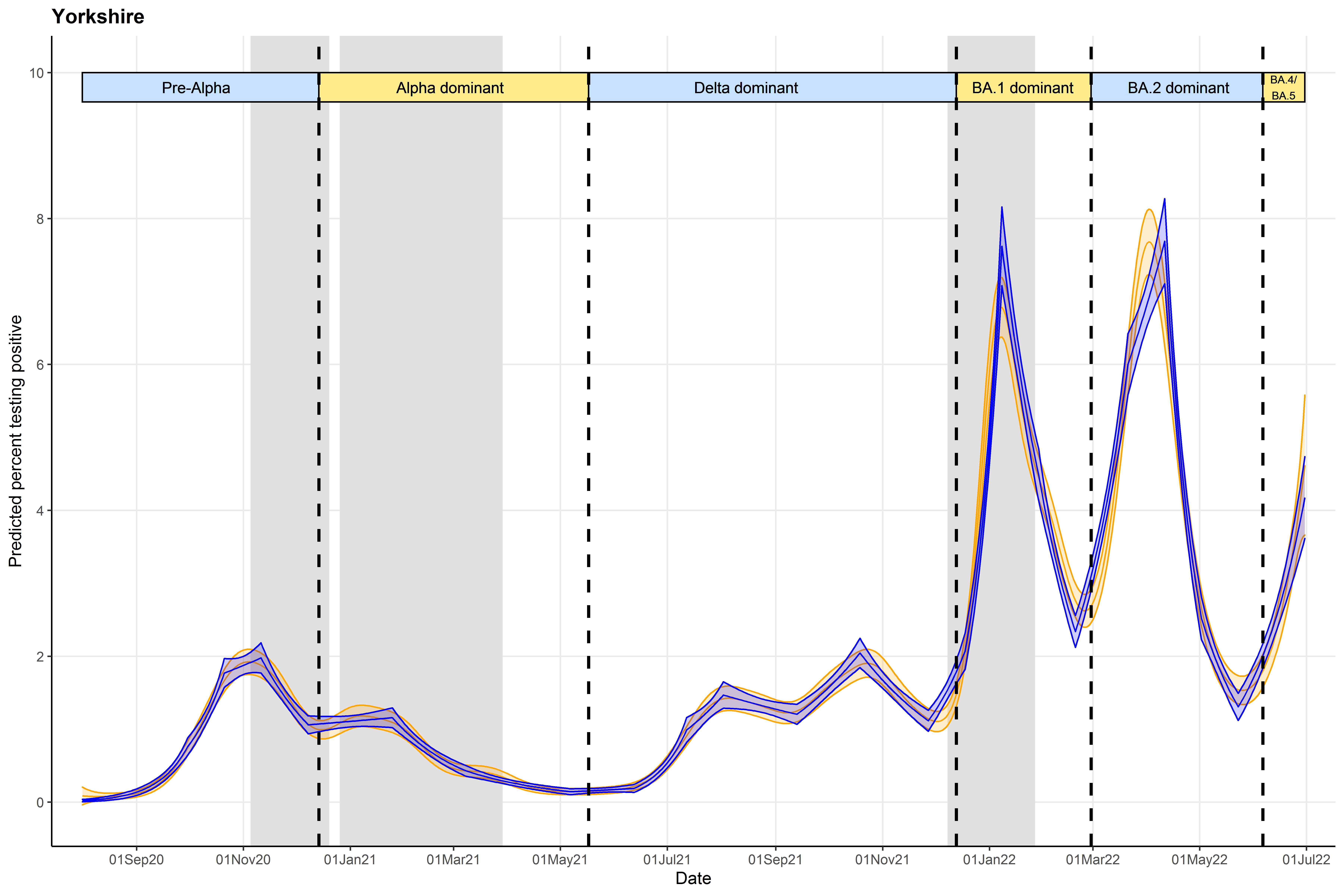

Note: Vertical dashed lines indicate periods when new variants became dominant, defined as >50% of positive swabs with cycle threshold (Ct)<30 being S-gene target positive (ORF1ab+N+S, ORF1ab+S, N+S gene positivity) in the Covid-19 Infection Survey for the pre-Alpha period (01 August 2020 - 13 December 2020), the Delta variant (17 May 2021 – 12 December 2021), and the Omicron BA.2 variant (28 February 2022 – 5 June 2022), and >50% Ct<30 S-gene target negative (ORF1ab+N gene positivity) for the Alpha variant (14 December 2020 – 16 May 2021), Omicron BA.1 variant (13 December 2021 – 27 February 2022), and Omicron BA4/BA.5 (6 June 2022 onwards) . Gray shaded indicate periods where stay/work from home laws were enforced, although specific restrictions varied across the time series.

#### Figure S5: Number of successive GAMs (zero to five), and ISR, finding the same change-points (top panel). Predicted positivity from final GAM for reference (bottom panel). Results are for Northern Ireland only.

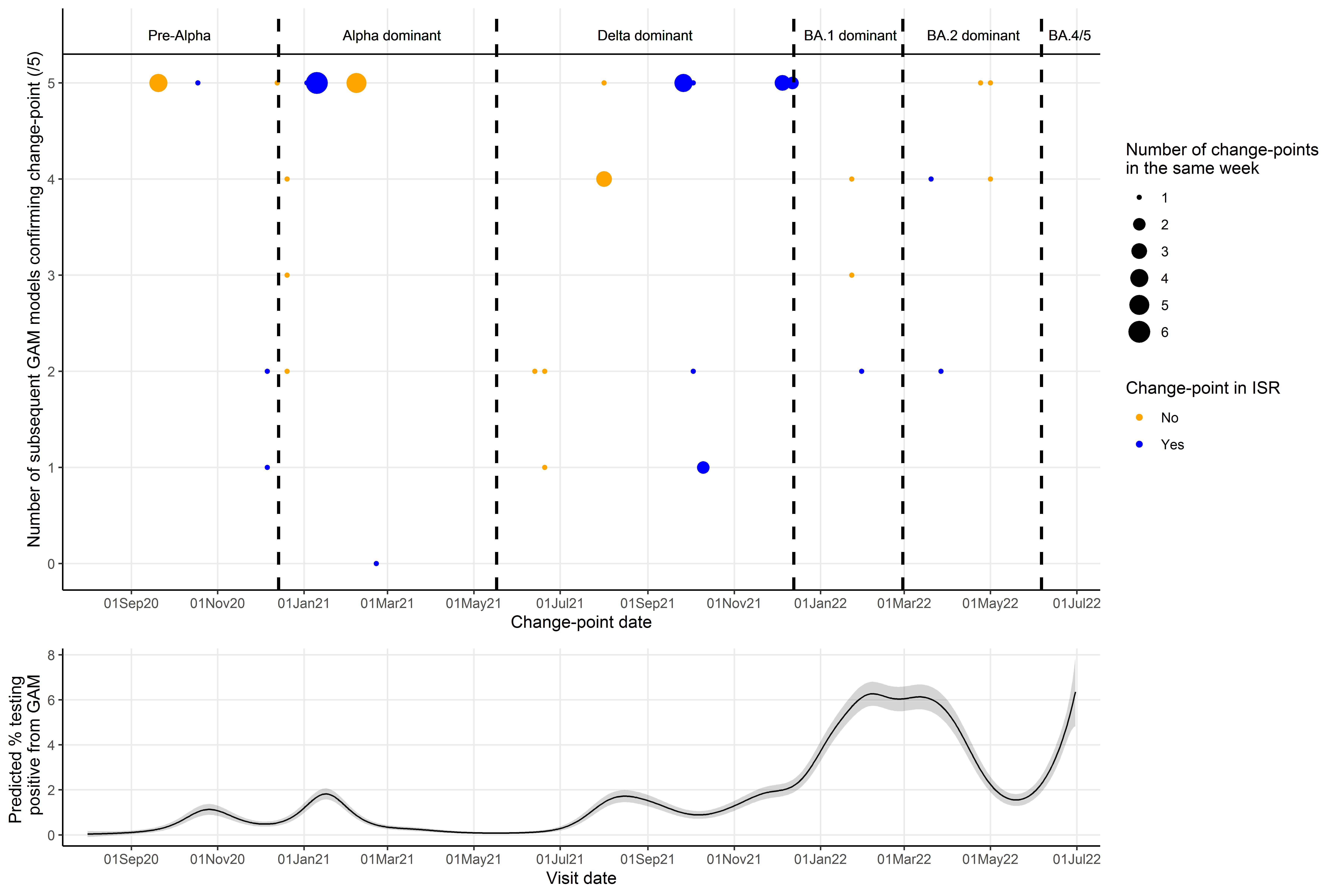

Note: Change-points in the same week (starting Monday) found in the same number of subsequent models were grouped together (indicated by size of circle). Points are blue if at least one change-point in that week was also found by ISR, and orange is no change-points in that week were found by ISR.

#### Figure S6: Predicted positivity (A), first derivatives (B), and second derivatives (C) estimated from GAMs fitted on the entire time-series for each geographical region, but only presented from 1st March to 30th June 2020. Original change-points based on the second derivative are shown in vertical blue lines, and additional change-points based on the first derivative are shown in vertical orange lines.

**A: Predicted positivity**

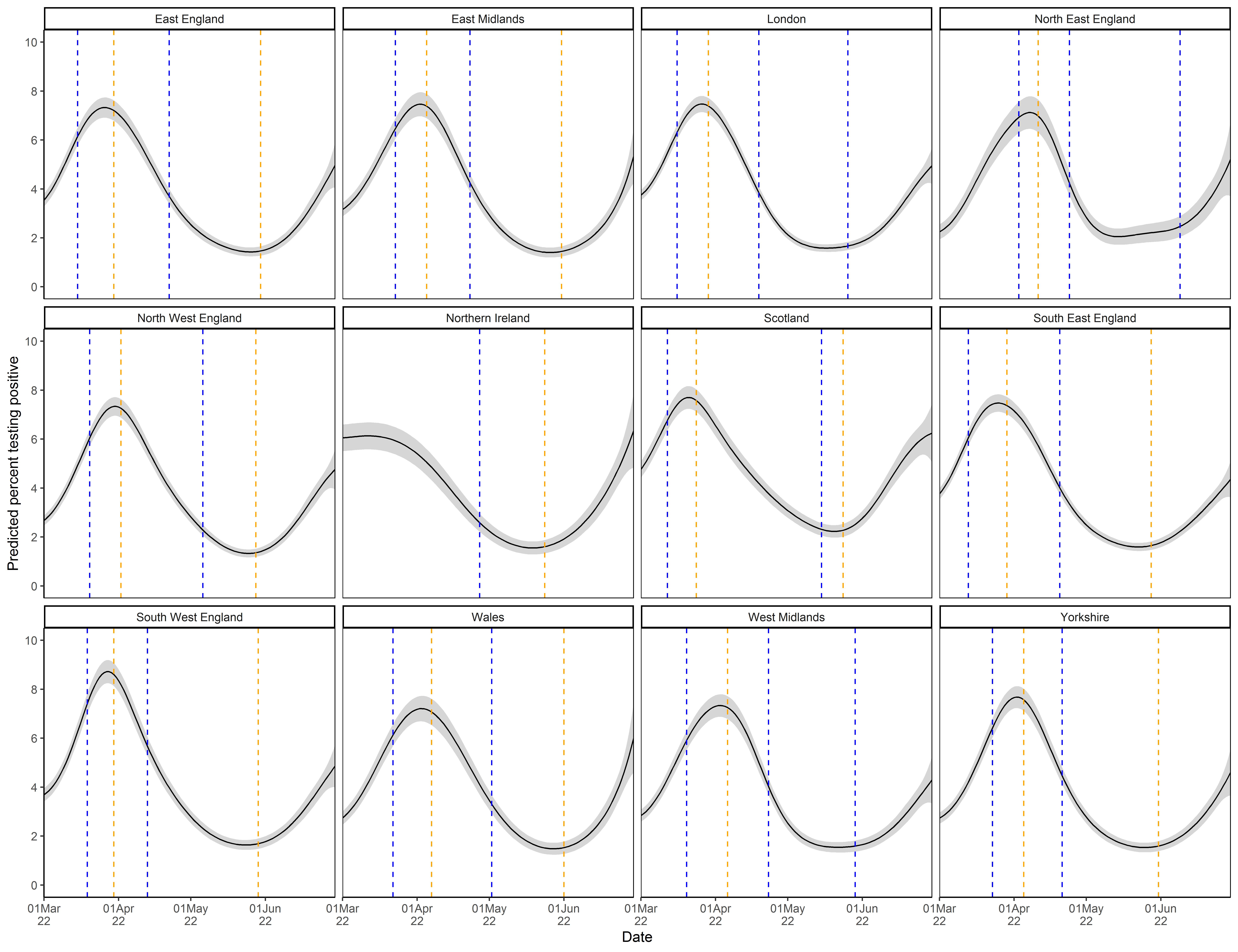

**B: First derivative**

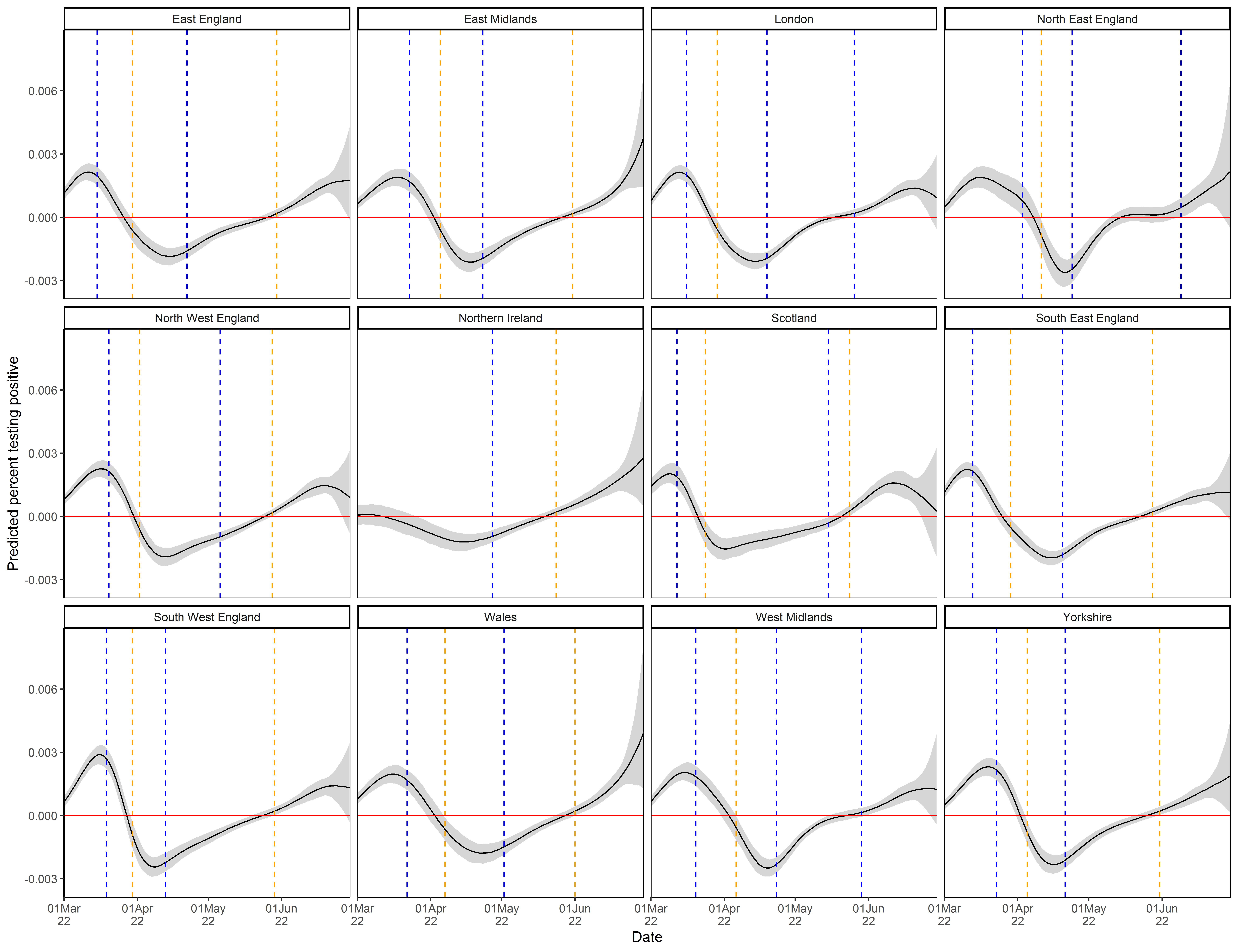

**C: Second derivative**

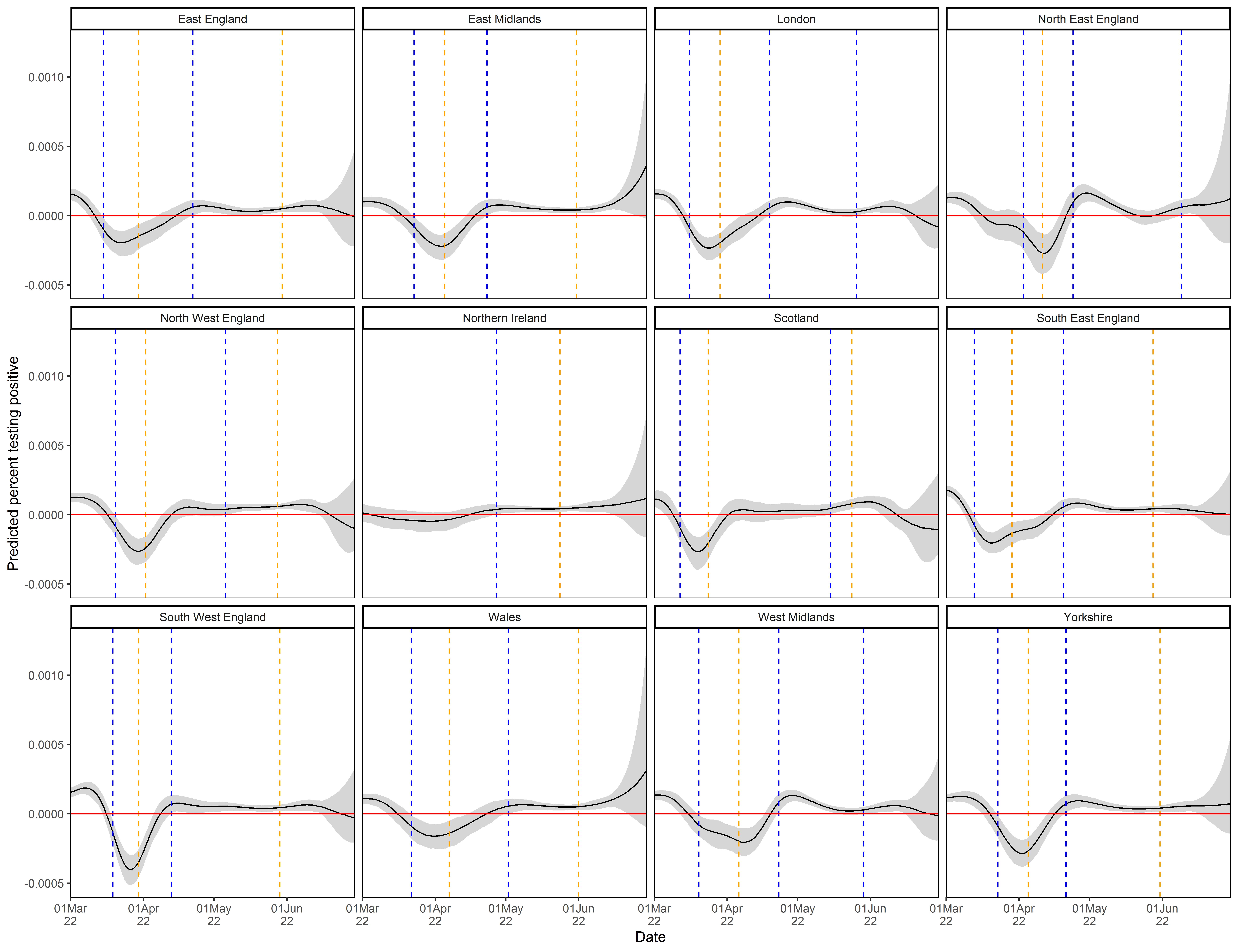

#### Figure S7: Predicted percentage testing positive for SARS-CoV-2 from ISR (blue) and GAMs (orange) for models run separately by age group for London

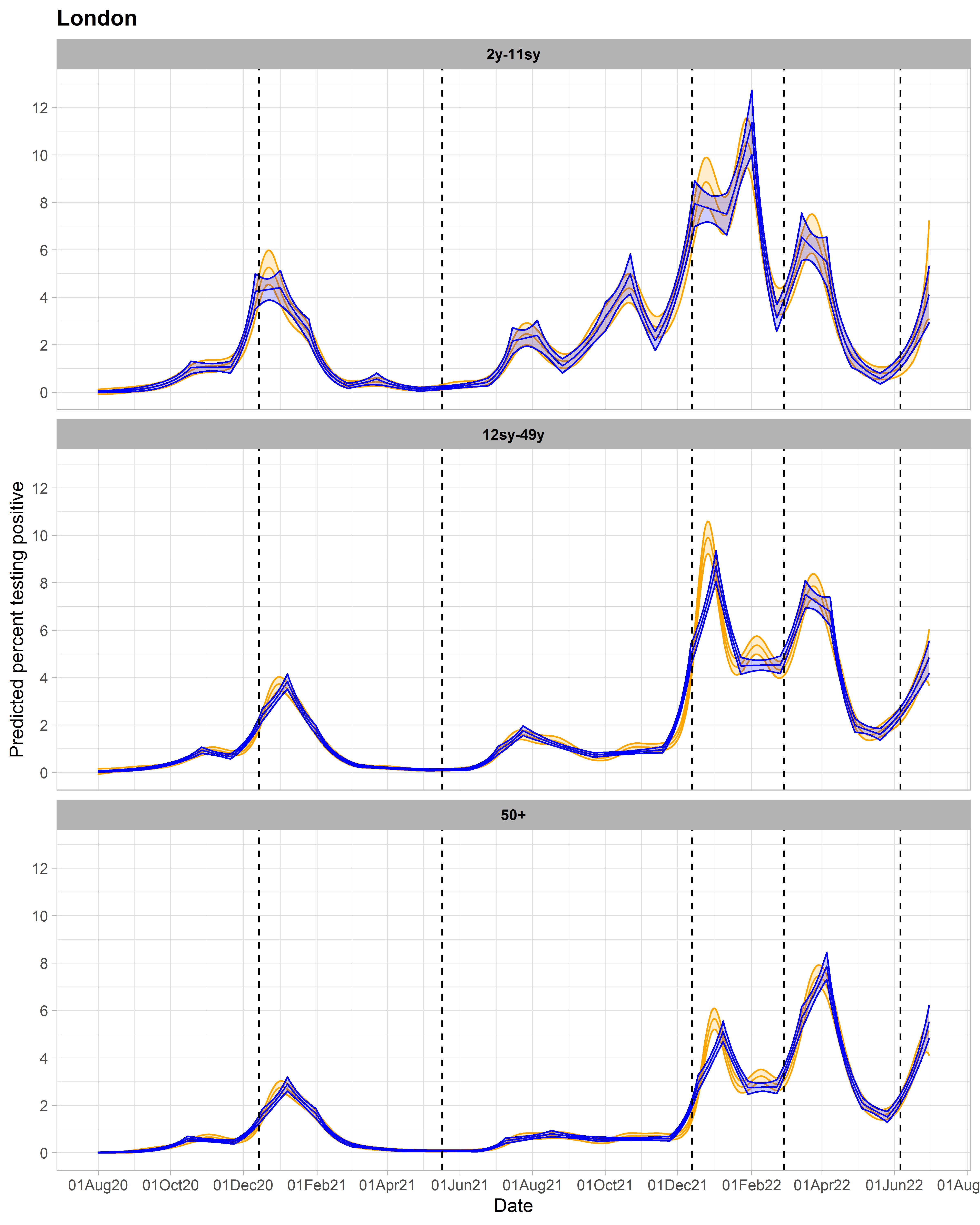

Note: Vertical dashed lines indicate periods when new variants became dominant, defined as >50% of positive swabs with cycle threshold (Ct)<30 being S-gene target positive (ORF1ab+N+S, ORF1ab+S, N+S gene positivity) in the Covid-19 Infection Survey for the pre-Alpha period (01 August 2020 - 13 December 2020), the Delta variant (17 May 2021 – 12 December 2021), and the Omicron BA.2 variant (28 February 2022 – 5 June 2022), and >50% Ct<30 S-gene target negative (ORF1ab+N gene positivity) for the Alpha variant (14 December 2020 – 16 May 2021), Omicron BA.1 variant (13 December 2021 – 27 February 2022), and Omicron BA4/BA.5 (6 June 2022 onwards).

#### Figure S8: Predicted percentage testing positive with first and second derivatives for different age groups in London.

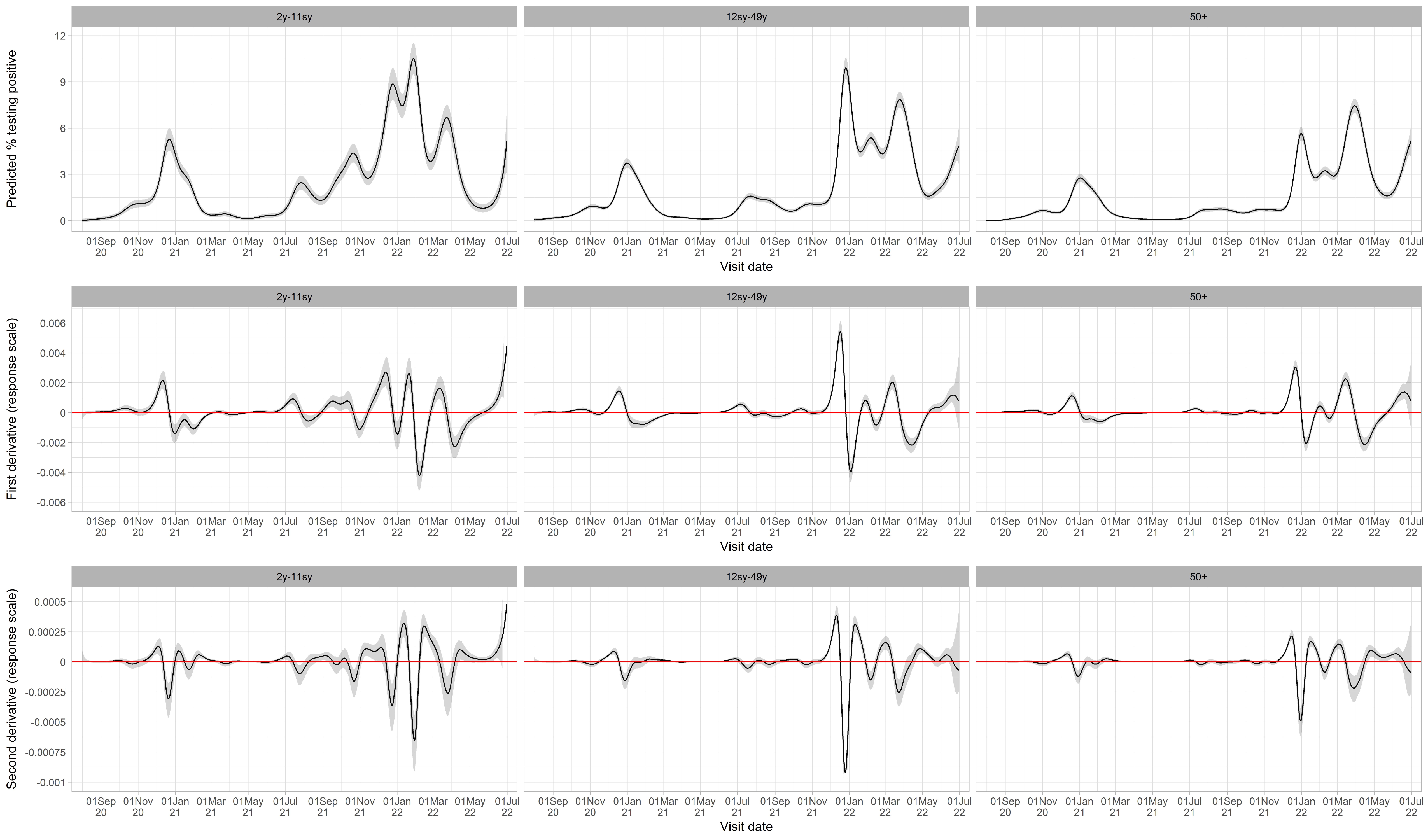

#### Figure S9: Raw daily percentage testing positive split by S-gene target positive and S-gene target failure.

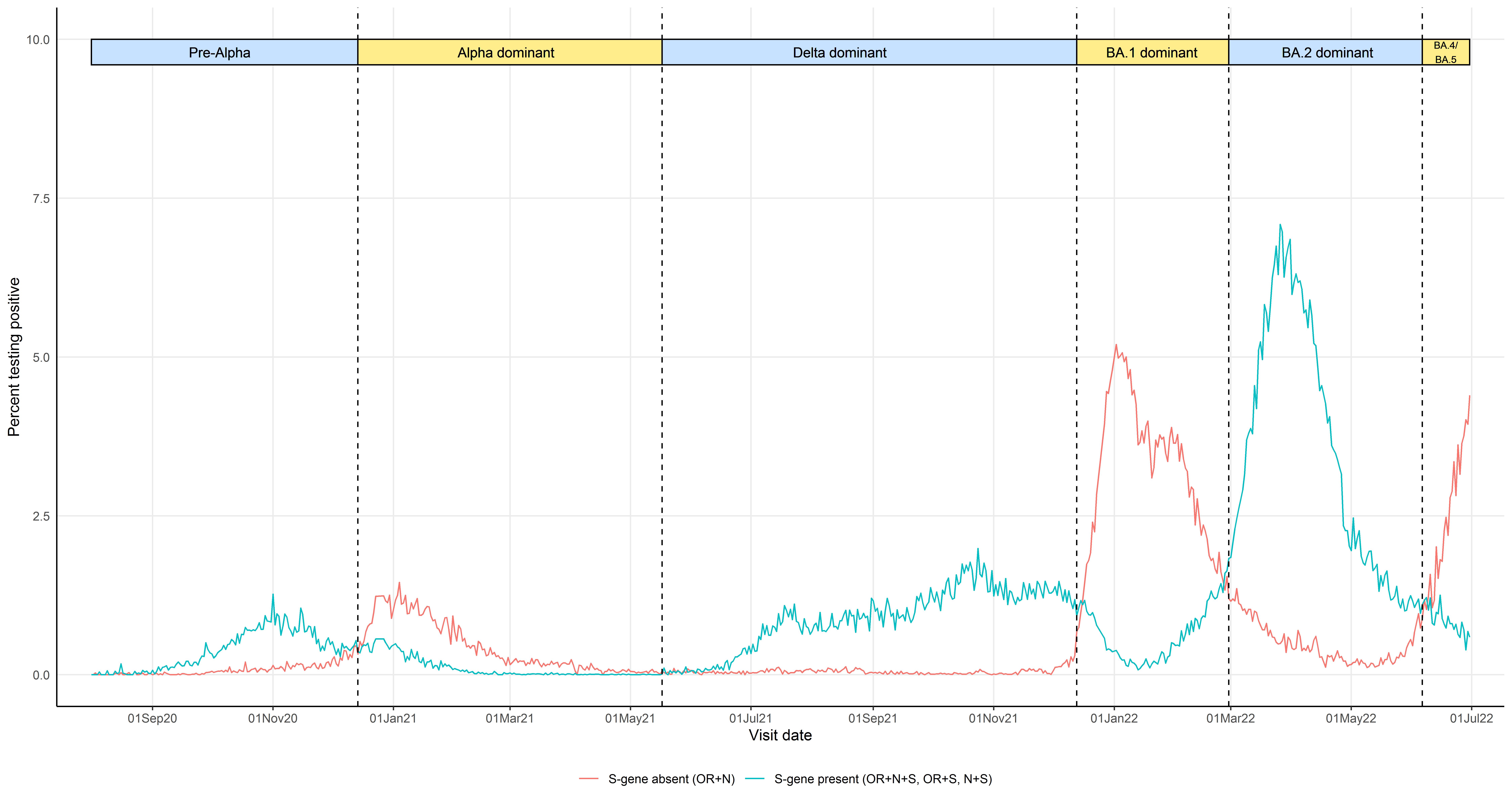

Note: Vertical dashed lines indicate periods when new variants became dominant, defined as >50% of positive swabs with cycle threshold (Ct)<30 being S-gene target positive (ORF1ab+N+S, ORF1ab+S, N+S gene positivity) in the Covid-19 Infection Survey for the pre-Alpha period (01 August 2020 - 13 December 2020), the Delta variant (17 May 2021 – 12 December 2021), and the Omicron BA.2 variant (28 February 2022 – 5 June 2022), and >50% Ct<30 S-gene target negative (ORF1ab+N gene positivity) for the Alpha variant (14 December 2020 – 16 May 2021), Omicron BA.1 variant (13 December 2021 – 27 February 2022), and Omicron BA4/BA.5 (6 June 2022 onwards).

#### Figure S10: Raw daily percentage testing positive split by S-gene target positive and S-gene target failure, by region.

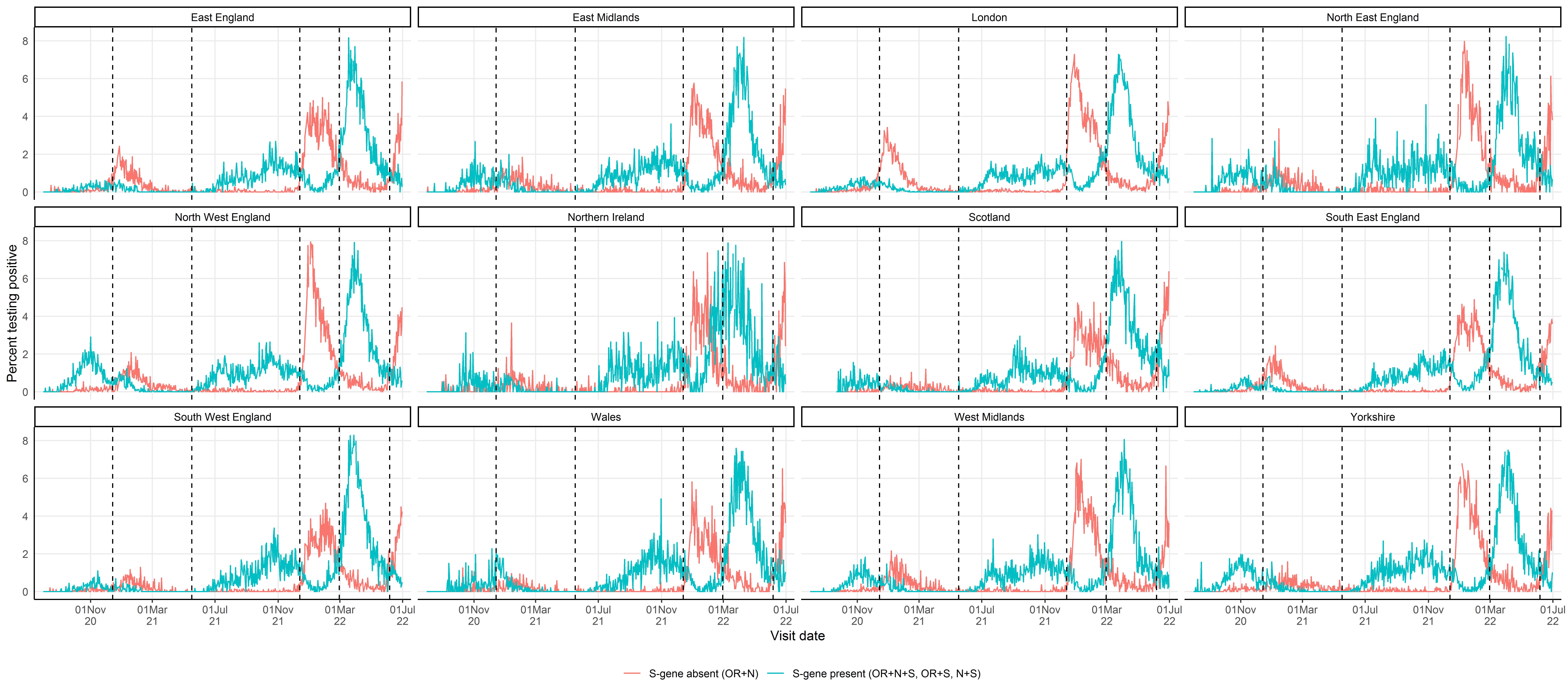

Note: Vertical dashed lines indicate periods when new variants became dominant, defined as >50% of positive swabs with cycle threshold (Ct)<30 being S-gene target positive (ORF1ab+N+S, ORF1ab+S, N+S gene positivity) in the Covid-19 Infection Survey for the pre-Alpha period (01 August 2020 - 13 December 2020), the Delta variant (17 May 2021 – 12 December 2021), and the Omicron BA.2 variant (28 February 2022 – 5 June 2022), and >50% Ct<30 S-gene target negative (ORF1ab+N gene positivity) for the Alpha variant (14 December 2020 – 16 May 2021), Omicron BA.1 variant (13 December 2021 – 27 February 2022), and Omicron BA4/BA.5 (6 June 2022 onwards).

#### Figure S11: Predicted daily percentage of visits testing positive for SARS-CoV-2 from ISR (blue) and GAMs (orange) for London only, split by SGTP and SGTF

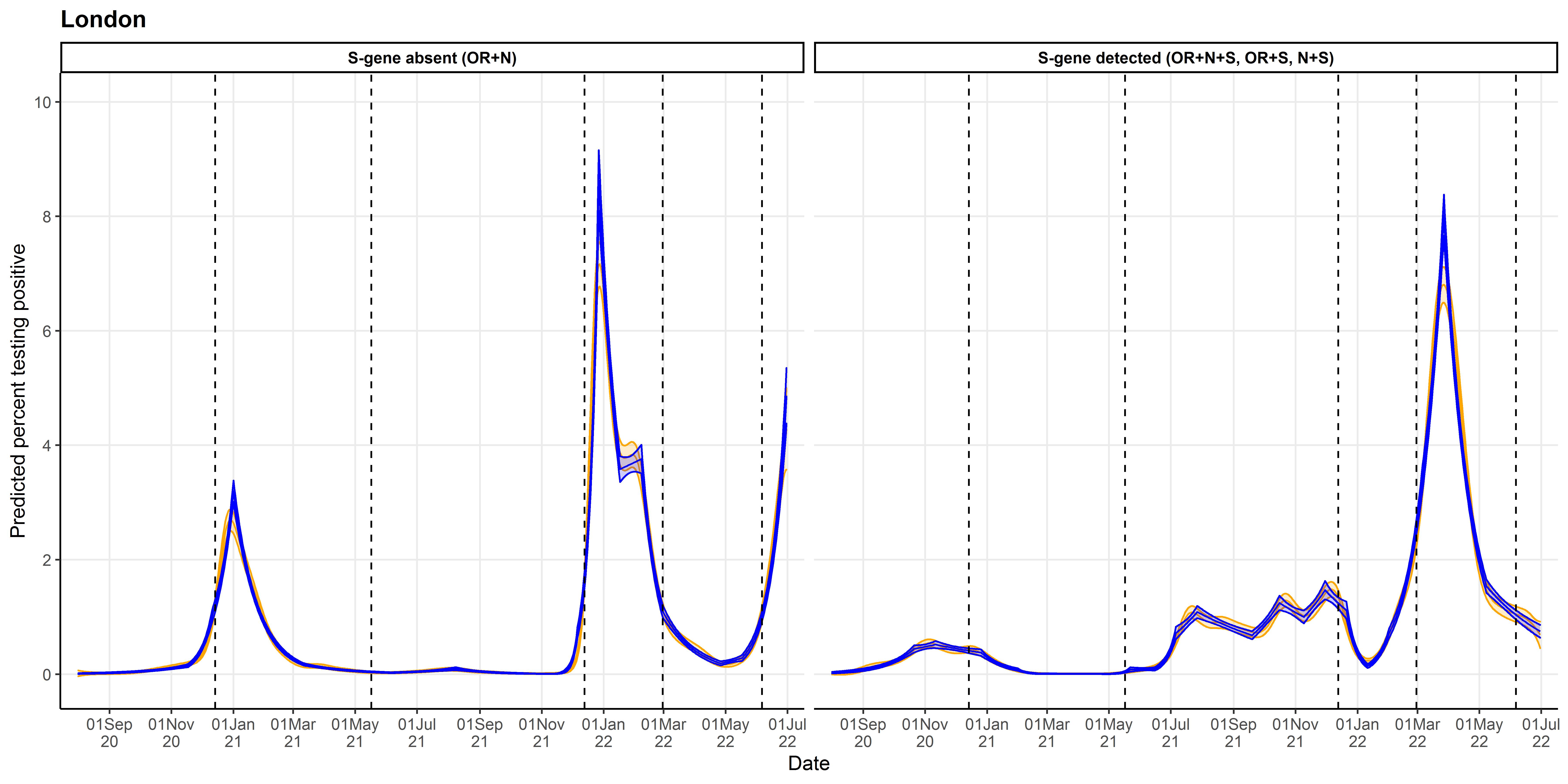

Note: Vertical dashed lines indicate periods when new variants became dominant, defined as >50% of positive swabs with cycle threshold (Ct)<30 being S-gene target positive (ORF1ab+N+S, ORF1ab+S, N+S gene positivity) in the Covid-19 Infection Survey for the pre-Alpha period (01 August 2020 - 13 December 2020), the Delta variant (17 May 2021 – 12 December 2021), and the Omicron BA.2 variant (28 February 2022 – 5 June 2022), and >50% Ct<30 S-gene target negative (ORF1ab+N gene positivity) for the Alpha variant (14 December 2020 – 16 May 2021), Omicron BA.1 variant (13 December 2021 – 27 February 2022), and Omicron BA4/BA.5 (6 June 2022 onwards).

### Supplementary Tables

#### Table S1: Characteristics of all visits included in analysis, split by swab result

| **Characteristic** | **Positive, n (%) or median (IQR)** | **Negative, n (%) or median (IQR)** | **Total, n (%) or median (IQR)** |
| --- | --- | --- | --- |
| **Age (years)** | 47 (27, 62) | 53 (34, 67) | 53 (34, 67) |
| **Age group** |  |  |  |
| 2y-11sy | 25,168 (17) | 977,775 (11) | 1,002,943 (11) |
| 12sy-49y | 54,950 (37) | 2,868,355 (33) | 2,923,305 (33) |
| 50+ | 67,160 (45) | 4,805,671 (55) | 4,872,831 (55) |
| **Sex** |  |  |  |
| Male | 70,733 (48) | 4,035,518 (46) | 4,106,251 (46) |
| Female | 76,545 (51) | 4,616,283 (53) | 4,692,828 (53) |
| **Geographical region** |  |  |  |
| Scotland | 10,679 (7) | 648,775 (7) | 659,454 (7) |
| North West England | 18,801 (12) | 994,441 (11) | 1,013,242 (11) |
| North East England | 5,831 (3) | 321,657 (3) | 327,488 (3) |
| Yorkshire | 12,889 (8) | 719,130 (8) | 732,019 (8) |
| East Midlands | 9,089 (6) | 547,238 (6) | 556,327 (6) |
| West Midlands | 11,042 (7) | 658,057 (7) | 669,099 (7) |
| East England | 12,834 (8) | 829,889 (9) | 842,723 (9) |
| Wales | 6,725 (4) | 428,267 (4) | 434,992 (4) |
| London | 26,286 (17) | 1,443,346 (16) | 1,469,632 (16) |
| South East England | 17,758 (12) | 1,128,635 (13) | 1,146,393 (13) |
| South West England | 10,823 (7) | 686,459 (7) | 697,282 (7) |
| Northern Ireland | 4,521 (3) | 245,907 (2) | 250,428 (2) |

#### Table S2: Characteristics of SARS-CoV-2 positive swabs, split by period in which different variants dominated

| **Characteristic** | **Pre-alpha** | **Alpha** | **Delta** | **Omicron BA.1** | **Omicron BA.2** | **Omicron BA.4/BA.5** |
| --- | --- | --- | --- | --- | --- | --- |
| **Date range** | 01 August 2020 to  13 December 2020 | 14 December 2020 to 16 May 2021 | 17 May 2021 to  12 December 2021 | 13 December 2021 to 27 February 2022 | 28 February 2022 to  05 June 2022 | 06 June 2022,  30 June 2022 |
| **Number of positives,**  **n (%)** | 12,263 (8) | 16,667 (11) | 26,805 (18) | 39,620 (27) | 45,318 (31) | 6,582 (4) |
| **Ct value, median (IQR)** | 28 (21, 32) | 30 (22, 33) | 25 (19, 31) | 24 (19, 30) | 24 (20, 30) | 23 (19, 28) |
| **Ct < 30, n (%)** | 7,203 (59) | 8,307 (50) | 19,107 (71) | 30,432 (77) | 34,200 (75) | 5,402 (82) |
| **SGTF**  **(% all pos) [% Ct <30]** | 705 (6) [10] | 6,593 (40) [79] | 242 (1) [1] | 25,462 (64) [84] | 3,206 (7) [9] | 4,054 (62) [75] |
| **S-gene detected**  **(% all pos) [% Ct <30]** | 6,452 (53) [90] | 1,669 (10) [20] | 18,844 (70) [99] | 4,925 (12) [16] | 30,985 (68) [91] | 1,345 (20) [25] |
| **Ct ≥ 30 (% all pos)** | 5,106 (42) | 8,405 (50) | 7,719 (29) | 9,233 (23) | 11,127 (25) | 1,183 (18) |

Note: Excluding 23 positives results without Ct values or genes detected available. Epochs were defined by >50% of positive swabs with cycle threshold (Ct)<30 being S-gene target positive (ORF1ab+N+S, ORF1ab+S, N+S gene positivity) in the Covid-19 Infection Survey for the pre-Alpha period (01 August 2020 - 13 December 2020), the Delta variant (17 May 2021 – 12 December 2021), and the Omicron BA.2 variant (28 February 2022 – 5 June 2022), and >50% Ct<30 S-gene target negative (ORF1ab+N gene positivity) for the Alpha variant (14 December 2020 – 16 May 2021), Omicron BA.1 variant (13 December 2021 – 27 February 2022), and Omicron BA4/BA.5 (6 June 2022 onwards) .

#### Table S3: Change-points corresponding to periods corresponding to emergence of four key SARS-CoVS-2 variants found by iterative sequential regression (ISR) and second derivatives of generalised additive models (GAM) for each geographical region, run on the full time-series, as shown in Figure 2.

| **Region** | **Coincident Variant** | **GAM breakpoint (DD.MM.YYYY)** | **ISR breakpoint**  **(DD.MM.YYYY)** | **ISR detection date** | **Days between ISR and GAM change-point*** | **Days between ISR change-point and detection** |
| --- | --- | --- | --- | --- | --- | --- |
| **East England** | Alpha | 20.11.2020 | 20.11.2020 | 14.12.2020 | 0 | 24 |
|  | Delta | 05.06.2021 | 09.06.2021 | 06.07.2021 | -4 | 27 |
|  | BA.1 | 27.11.2021 | 24.11.2021 | 18.12.2021 | 3 | 24 |
|  | BA.2 | 14.02.2022 | 16.02.2022 | 12.03.2022 | -2 | 24 |
| **East Midlands** | Alpha | **Not found** | 11.12.2020 | 04.01.2021 | n/a | 24 |
|  | Delta | 01.06.2021 | 12.06.2021 | 06.07.2021 | -11 | 24 |
|  | BA.1 | 28.11.2021 | 03.12.2021 | 27.12.2021 | -5 | 24 |
|  | BA.2 | 15.02.2022 | 16.02.2022 | 12.03.2022 | -1 | 24 |
| **London** | Alpha | 20.11.2020 | 26.11.2020 | 20.12.2020 | -6 | 24 |
|  | Delta | 09.06.2021 | 06.06.2021 | 30.06.2021 | 3 | 24 |
|  | BA.1 | 24.11.2021 | 30.11.2021 | 24.12.2021 | -6 | 24 |
|  | BA.2 | 15.02.2022 | 28.02.2022 | 24.03.2022 | -13 | 24 |
| **North East** | Alpha | 06.12.2020 | 08.12.2020 | 01.01.2021 | -2 | 24 |
|  | Delta | 25.05.2021 | 06.06.2021 | 09.07.2021 | -12 | 33 |
|  | BA.1 | 04.12.2021 | 06.12.2021 | 30.12.2021 | -2 | 24 |
|  | BA.2 | 15.02.2022 | 16.02.2022 | 12.03.2022 | -1 | 24 |
| **Northern Ireland** | Alpha | 19.11.2020 | 05.12.2020 | 04.01.2021 | -16 | 30 |
|  | Delta | 09.06.2021 | 09.06.2021 | 03.07.2021 | 0 | 24 |
|  | BA.1 | 07.12.2021 | 12.12.2021 | 05.01.2022 | -5 | 24 |
|  | BA.2 | n/a | n/a | n/a | n/a | n/a |
| **North West** | Alpha | 01.12.2020 | 05.12.2020 | 29.12.2020 | -4 | 24 |
|  | Delta | 14.04.2021 | 01.05.2021 | 25.05.2021 | -17 | 24 |
|  | BA.1 | 30.11.2021 | 24.11.2021 | 18.12.2021 | 6 | 24 |
|  | BA.2 | 12.02.2022 | 16.02.2022 | 12.03.2022 | -4 | 24 |
| **Scotland** | Alpha | **Not found** | 20.11.2020 | 23.12.2020 | n/a | 33 |
|  | Delta | 26.05.2021 | 03.06.2021 | 27.06.2021 | -8 | 24 |
|  | BA.1 | 01.12.2021 | 30.11.2021 | 24.12.2021 | 1 | 24 |
|  | BA.2 | 15.01.2022 | 01.02.2022 | 25.02.2022 | -17 | 24 |
| **South East** | Alpha | 23.11.2020 | 26.11.2020 | 20.12.2020 | -3 | 24 |
|  | Delta | 08.06.2021 | 21.06.2021 | 15.07.2021 | -13 | 24 |
|  | BA.1 | 05.12.2021 | 03.12.2021 | 27.12.2021 | 2 | 24 |
|  | BA.2 | 15.02.2022 | 19.02.2022 | 15.03.2022 | -4 | 24 |
| **South West** | Alpha | 26.11.2020 | 08.12.2020 | 01.01.2021 | -12 | 24 |
|  | Delta | 23.05.2021 | 01.05.2021 | 25.05.2021 | 22 | 24 |
|  | BA.1 | 06.11.2021 | 21.11.2021 | 18.12.2021 | -15 | 27 |
|  | BA.2 | 19.02.2022 | 28.02.2022 | 24.03.2022 | -9 | 24 |
| **Wales** | Alpha | 14.11.2020 | 20.11.2020 | 14.12.2020 | -6 | 24 |
|  | Delta | 01.06.2021 | 27.06.2021 | 01.10.2021 | -26 | 96 |
|  | BA.1 | 01.12.2021 | 06.12.2021 | 30.12.2021 | -5 | 24 |
|  | BA.2 | 14.02.2022 | 16.02.2022 | 12.03.2022 | -2 | 24 |
| **West Midlands** | Alpha | 30.11.2020 | 05.12.2020 | 29.12.2020 | -5 | 24 |
|  | Delta | 25.05.2021 | 10.05.2021 | 06.06.2021 | 15 | 27 |
|  | BA.1 | 28.11.2021 | 27.11.2021 | 21.12.2021 | 1 | 24 |
|  | BA.2 | 15.02.2022 | 13.02.2022 | 09.03.2022 | 2 | 24 |
| **Yorkshire** | Alpha | 03.12.2020 | 08.12.2020 | 01.01.2021 | -5 | 24 |
|  | Delta | 09.06.2021 | 12.06.2021 | 06.07.2021 | -3 | 24 |
|  | BA.1 | 27.11.2021 | 27.11.2021 | 21.12.2021 | 0 | 24 |
|  | BA.2 | 14.02.2022 | 19.02.2022 | 15.03.2022 | -5 | 24 |

*Negative values indicate earlier occurrence of change-points using GAMs, compared with ISR.

#### Table S4: Comparison of change-points detected by generalised additive models run on the full time series from 1st August 2020, 16-week, 24-week, and 32-week periods for London

| **Model end date (days from 1^st^ August 2020)** | **Change-point dates from model run from 1^st^ August 2020 [duration*, days]** | **Change-point dates (days between current change-point and change-point in full model) [duration*, days]** | | |
| --- | --- | --- | --- | --- |
|  |  | **16-week model (112 days)** | **24-week model (168 days)** | **32-week model (224 days)** |
| 26.11.2020 (118 days) | 06.11.2020 [14] | 04.11.2020 (-2) [17] | n/a | n/a |
| 21.01.2021 (174 days) | 09.12.2020 [5] | - | 11.12.2020 (2) [3] | n/a |
|  | 23.12.2020 [9] | 25.12.2020 (2) [6] | 24.12.2020 (1) [7] | n/a |
|  | - | 05.01.2021 (n/a) [3] | - | n/a |
| 18.03.2021 (230 days) | 10.02.2021 [1] | - | - | 10.02.2021 (0) [3] |
|  | 12.02.2021 [2] | - | - | 14.02.2021 (-2) [1] |
|  | - | - | - | 21.02.2021 (n/a) [2] |
|  | 23.02.2021 [9] | 26.02.2021 (3) [7] | - | 24.02.2021 (-1) [9] |
| 13.05.2021 (286 days) | No change-points | No change-points | No change-points | No change-points |
| 08.07.2021 (342 days) | 13.06.2021 [17] | - | - | 14.06.2021 (-1) [19] |
| 02.09.2021 (398 days) | 13.07.2021 [12] | 14.07.2021 (1) [9] | 14.07.2021 (1) [11] | 14.07.2021 (1) [12] |
| 28.10.2021 (454 days) | - | 10.09.2021 (n/a) [45] | - | - |
|  | 04.10.2021 [6] | - | - | 04.10.2021 (0) [5] |
| 23.12.2021 (510 days) | 04.11.2021 [10] | - | - | - |
|  | 27.11.2021 [27] | 04.12.2021 (7) [12] | 03.12.2021 (6) [14] | 02.12.2021 (5) [16] |
| 17.02.2022 (566 days) | 06.01.2022 [18] | 12.01.2022 (6) [6] | 12.01.2022 (6) [5] | 11.01.2022 (5) [8] |
|  | 02.02.2022 [5] | - | - | - |

*Duration = number of days which the credible interval of the second derivative did not contain zero

Note: Change-points recorded as “n/a” are not applicable as the 24-week and/or 32-week model is identical to the model run from 1^st^ August 2020.

#### Table S5: Detection dates for GAMs and ISR for London and Northern Ireland

| **Geographical region** | **Change-point in final GAM** | **GAM detection date** | **ISR change-point** | **ISR detection date** | **Difference in GAM change-point & detection** | **Difference in ISR change-point & detection** | **Difference in GAM & ISR change-points** | **Difference in GAM & ISR detection** |
| --- | --- | --- | --- | --- | --- | --- | --- | --- |
| **London** | 26.09.2020 | 22.10.2020 | n/a | n/a | 26 | n/a | n/a | n/a |
|  | n/a | n/a | 15.10.2020 | 08.11.2020 | n/a | 24 | n/a | n/a |
|  | 02.11.2020 | 19.11.2020 | 05.11.2020 | 29.11.2020 | 17 | 24 | -3 | -10 |
|  | 20.11.2020 | 03.12.2020 | 26.11.2020 | 20.12.2020 | 13 | 24 | -6 | -17 |
|  | 19.12.2020 | 07.01.2021 | 17.12.2020 | 10.01.2021 | 19 | 24 | 2 | -3 |
|  | n/a | n/a | 07.01.2021 | 31.01.2021 | n/a | 24 | n/a | n/a |
|  | 23.01.2021 | n/a | 28.01.2021 | 21.02.2021 | n/a | 24 | -5 | n/a |
|  | 05.02.2021 | 25.02.2021 | n/a | n/a | 20 | n/a | n/a | n/a |
|  | n/a | n/a | 02.03.2021 | 26.03.2021 | n/a | 24 | n/a | n/a |
|  | n/a | n/a | 01.05.2021 | 25.05.2021 | n/a | 24 | n/a | n/a |
|  | 09.06.2021 | 01.07.2021 | 06.06.2021 | 30.06.2021 | 22 | 24 | 3 | 1 |
|  | 12.07.2021 | 29.07.2021 | 06.07.2021 | 30.07.2021 | 17 | 24 | 6 | -1 |
|  | n.a | n/a | 27.07.2021 | 20.08.2021 | n/a | 24 | n/a | n/a |
|  | 25.09.2021 | 21.10.2021 | 19.09.2021 | 13.10.2021 | 26 | 24 | 6 | 8 |
|  | 15.10.2021 | 04.11.2021 | 16.10.2021 | 09.11.2021 | 20 | 24 | -1 | -5 |
|  | 01.11.2021 | 25.11.2021 | n/a | n/a | 24 | n/a | n/a | n/a |
|  | n/a | n/a | 09.11.2021 | 03.12.2021 | n/a | 24 | NA | NA |
|  | 24.11.2021 | n/a | 30.11.2021 | 24.12.2021 | NA | 24 | -6 | NA |
|  | 20.12.2021 | 30.12.2021 | 21.12.2021 | 14.01.2022 | 10 | 24 | -1 | -15 |
|  | 06.01.2022 | 27.01.2022 | 11.01.2022 | 04.02.2022 | 21 | 24 | -5 | -8 |
|  | 29.01.2022 | n/a | n/a | n/a | n/a | n/a | n/a | n/a |
|  | n/a | n/a | 07/02.2022 | 03.03.2022 | n/a | 24 | n/a | n/a |
|  | 15.02.2022 | 23.06.2022 | n/a | n/a | 128 | n/a | n/a | n/a |
|  | n/a | n/a | 28.02.2022 | 24.03.2022 | n/a | 24 | n/a | n/a |
|  | 16.03.2022 | 19.05.2022 | 21.03.2022 | 14.04.2022 | 64 | 24 | -5 | 35 |
|  | n/a | n/a | 11.04.2022 | 05.05.2022 | n/a | 24 | n/a | n/a |
|  | 19.04.2022 | 12.05.2022 | n/a | n/a | 23 | NA | n/a | n/a |
|  | n/a | n/a | 02.05.2022 | 26.05.2022 | n/a | 24 | n/a | n/a |
|  | 26.05.2022 | n/a | 23.05.2022 | 16.06.2022 | n/a | 24 | 3 | n/a |
| **Northern Ireland** | 14.09.2020 | 31.12.2020 | n/a | n/a | 108 | n/a | n/a | n/a |
|  | 17.10.2020 | 10.12.2020 | 15.10.2020 | 08.11.2020 | 54 | 24 | 2 | 32 |
|  | 19.11.2020 | n/a | n/a | n/a | n/a | n/a | n/a | n/a |
|  | n/a | n/a | 05.12.2020 | 04.01.2021 | n/a | 30 | n/a | n/a |
|  | 07.01.2021 | 21.01.2021 | 07.01.2021 | 31.01.2021 | 14 | 24 | 0 | -10 |
|  | 04.02.2021 | 11.03.2021 | 28.01.2021 | 21.02.2021 | 35 | 24 | 7 | 18 |
|  | n/a | n/a | 27.02.2021 | 23.03.2021 | n/a | 24 | n/a | n/a |
|  | 09.06.2021 | 12.08.2021 | 09.06.2021 | 03.07.2021 | 64 | 24 | n/a | 40 |
|  | n/a | n/a | 21.07.2021 | 14.08.2021 | n/a | 24 | n/a | n/a |
|  | 02.08.2021 | 26.08.2021 | n/a | n/a | 24 | n/a | n/a | n/a |
|  | n/a | n/a | 20.08.2021 | 01.10.2021 | n/a | 42 | n/a | n/a |
|  | 28.09.2021 | 28.10.2021 | 04.10.2021 | 28.10.2021 | 30 | 24 | -6 | 0 |
|  | 07.12.2021 | 06.01.2022 | 12.12.2021 | 05.01.2022 | 30 | 24 | -5 | 1 |
|  | n/a | n/a | 02.01.2022 | 13.02.2022 | n/a | 42 | n/a | n/a |
|  | n/a | n/a | 04.02.2022 | 28.02.2022 | n/a | 24 | n/a | n/a |
|  | n/a | n/a | 24.03.2022 | 17.04.2022 | n/a | 24 | n/a | n/a |
|  | 27.04.2022 | 05.05.2022 | n/a | n/a | 8 | n/a | n/a | n/a |
|  | n/a | n/a | 14.05.2022 | 07.06.2022 | n/a | 24 | n/a | n/a |

#### Table S6: Change-points from GAMs and ISR fitted on the full time-series for BA.4/ BA.5. Change-points from GAMs are presented for both change-points defined by the second derivative alone, as well as additional change-points based on the first derivative.

| **Geographical region** | **GAM change-point estimated using second derivative only** | **GAM change-point incorporating additional change-points from first derivative** | **ISR change-point** |
| --- | --- | --- | --- |
| **East England** | Not found | 30.05.2022 | 23.05.2022 |
| **East Midlands** | Not found | 31.05.2022 | 01.06.2022 |
| **London** | 26.05.2022 | No change | 23.05.2022 |
| **North East** | 09.06.2022 | No change | 11.05.2022 |
| **Northern Ireland** | Not found | 24.05.2022 | 14.05.2022 |
| **North West** | Not found | 28.05.2022 | 01.06.2022 |
| **Scotland** | Not found | 24.05.2022 | 29.05.2022 |
| **South East** | Not found | 28.05.2022 | 20.05.2022 |
| **South West** | Not found | 29.05.2022 | 23.05.2022 |
| **Wales** | Not found | 01.06.2022 | 17.05.2022 |
| **West Midlands** | 29.05.2022 | No change | 01.06.2022 |
| **Yorkshire** | Not found | 31.05.2022 | 23.05.2022 |

#### Table S7: Change-points from GAMs (run on the full time-series) and ISR run separately by age, separately by S gene detection and overall in London.

| **Variant** | **Sub-Group** | **GAM change-point date** | **ISR change-point date** | **ISR detection date** | **Days between GAM and ISR change-points*** | **Days between ISR change-point and detection date** |
| --- | --- | --- | --- | --- | --- | --- |
| **Alpha** | 2y-11sy | 19.11.2020 | 20.11.2020 | 14.12.2020 | -1 | 24 |
|  | 12sy-49 | 20.11.2020 | 20.11.2020 | 14.12.2020 | 0 | 24 |
|  | 50+ | 20.11.2020 | 23.11.2020 | 17.12.2020 | -3 | 24 |
|  | S-gene absent | 19.11.2020 | 17.11.2020 | 11.12.2020 | 2 | 24 |
|  | **Overall model** | **20.11.2020** | **26.11.2020** | 20.12.2020 | -6 | 24 |
| **Delta** | 2y-11sy | 16.06.2021 | 24.06.2021 | 21.07.2021 | -8 | 27 |
|  | 12sy-49 | 05.06.2021 | 06.06.2021 | 30.06.2021 | -1 | 24 |
|  | 50+ | 15.06.2021 | 15.06.2021 | 09.07.2021 | 0 | 24 |
|  | S-gene detected | 10.06.2021 | 15.06.2021 | 09.07.2021 | -5 | 24 |
|  | **Overall model** | **09.06.2021** | **06.06.2021** | 30.06.2021 | 3 | 24 |
| **Omicron BA.1** | 2y-11sy | 05.11.2021 | 12.11.2021 | 06.12.2021 | -7 | 24 |
|  | 12sy-49 | 19.11.2021 | 18.11.2021 | 12.12.2021 | 1 | 24 |
|  | 50+ | 25.11.2021 | 24.11.2021 | 18.12.2021 | 1 | 24 |
|  | S-gene absent | 11.11.2021 | 12.11.2021 | 09.12.2021 | -1 | 27 |
|  | **Overall model** | **24.11.2021** | **30.11.2021** | 24.12.2021 | -6 | 24 |
| **Omicron BA.2** | 2y-11sy | 09.02.2022 | 22.02.2022 | 18.03.2022 | -13 | 24 |
|  | 12sy-49y | 18.02.2022 | 25.02.2022 | 21.03.2022 | -7 | 24 |
|  | 50y+ | 19.02.2022 | 22.02.2022 | 18.03.2022 | -3 | 24 |
|  | S-gene detected | 17.02.2022 | 11.01.2022 | 04.02.2022 | 37 | 24 |
|  | **Overall model** | **15.02.2022** | **28.02.2022** | 24.03.2022 | -13 | 24 |

Note:. Overall estimates from GAMs and ISR include all age groups with the outcome of all positives. *Negative values indicate earlier occurrence of change-points using GAMs, compared with ISR
